# Improved non-invasive detection of ictal and interictal epileptiform activity using Optically Pumped Magnetometers

**DOI:** 10.1101/2022.11.03.22281836

**Authors:** Arjan Hillebrand, Niall Holmes, Ndedi Sijsma, George C. O’Neill, Tim M. Tierney, Niels Liberton, Anine H. Stam, Nicole van Klink, Cornelis J. Stam, Richard Bowtell, Matthew J. Brookes, Gareth R. Barnes

## Abstract

Magneto- and Electroencephalography (MEG/EEG) are important techniques for the diagnosis and pre-surgical evaluation of epilepsy. Yet, in current cryogen-based MEG systems the sensors are offset from the scalp, which limits the signal-to-noise ratio (SNR) and thereby the sensitivity to activity from deep structures such as the hippocampus. This effect is amplified in children, for whom adult-sized fixed-helmet systems are typically too big. Moreover, ictal recordings with fixed-helmet systems are problematic because of limited movement tolerance. Optically Pumped Magnetometers (OPMs) can be placed directly on the scalp, thereby improving SNR and consequently the sensitivity to, and localisation accuracy of, epileptiform activity. In addition, recording during seizures becomes feasible with these wearable sensors.

We aimed to demonstrate these advantages of OPMs in a clinical population. Three adults with known weak sources of interictal epileptiform discharges (IEDs), along with three children with focal epilepsy and one adult with frequent seizures underwent MEG recordings using a 12-channel OPM-system and a 306-channel cryogen-based whole-head system. Performance of the two systems was compared in terms of IED-rate and SNR.

In one patient the OPMs detected IEDs that were not found with the SQUID-system. In one patient the spike yield was higher for the OPM data (9.00 versus 6.76), with negligible difference in SNR compared to the SQUID data (3.85 versus 3.93; U = -2.86, d = -0.14). This was also the case for a patient with a spike yield that was comparable to that for the SQUID data (after accounting for unilateral coverage with the OPMs; SNR 4.47 versus 4.57; U = -3.81, d = -0.14). For one patient the spike yield (11.03 versus 24.50) and SNR (4.39 versus 4.05; U = 9.53, d = -0.36) were both lower for the OPMs. In two patients no IEDs were found with either system. Importantly, the wearability of OPMs enabled the recording of seizure activity in a patient with hyperkinetic movements during the seizure. The observed ictal onset and semiology were in agreement with previous video- and stereo-EEG recordings.

Overall, OPM data were very much comparable to those obtained with a cryogenic system: OPMs outperformed SQUIDs for two of the four patients with IEDs, with either a higher spike yield, or an ability to detect IEDs that were not observable in the SQUID data. For three patients the SNRs of IEDs were (slightly) lower in the OPM data than in the SQUID data, but with negligible effect sizes for two of these patients. The relatively cheap technology, in combination with reduced running and maintenance costs, means that OPM-based MEG could be used more widely than current MEG systems, and may become an affordable alternative to scalp EEG, with the potential benefits of increased spatial accuracy, reduced sensitivity to volume conduction/field spread, and increased sensitivity to deep sources. Wearable MEG thus provides an unprecedented opportunity for epilepsy, and given its patient-friendliness, we envisage that it will not only be used for presurgical evaluation of epilepsy patients, but also for diagnosis after a first seizure.

## 1. Introduction

Magneto- and Electroencephalography (MEG/EEG) are important techniques for the diagnosis (Colon et al., 2009) and pre-surgical evaluation (Nissen et al., 2016; Rampp et al., 2019) of epilepsy. For patients with focal refractory epilepsy, seizure freedom can be achieved through epilepsy surgery by removing the epileptogenic zone (EZ), which is defined as the area of cortex that is necessary and sufficient for initiating seizures and whose removal (or disconnection) is necessary for complete abolition of seizures (Luders et al., 2006). This requires the generation of a hypothesis about the location of the EZ during the pre-surgical workup using measurements from non-invasive techniques such as MEG/EEG, or invasive recordings using intracranial electrodes (Shah and Mittal, 2014). Interictal epileptiform discharges (IEDs) and ictal activity as identified in presurgical MEG/EEG help to identify the irritative zone (the area of cortical tissue that generates IEDs) and the seizure onset zone (the area of cortex from which clinical seizures are generated), respectively, both of which may overlap with the EZ (Jehi, 2018; Rosenow and Luders, 2001). The pre-surgical workup, and thereby surgery, needs to be improved though, as seizure freedom is currently achieved in only two-thirds of the patients who undergo surgery (Najm et al., 2013; Tellez-Zenteno and Wiebe, 2008).

Current techniques have their limitations: invasive EEG is burdensome to the patient, expensive, has risk of complications, and may still be inconclusive (Bekelis et al., 2013). Clinical scalp-EEG recordings have limited spatial resolution, which can be mitigated to some extent with high-density recordings in combination with advanced head- and source-modelling (Mégevand and Seeck, 2020; Nemtsas et al., 2017). Although MEG generally has a higher spatial resolution, its sensitivity to activity from deep structures could improve when recordings with higher signal-to-noise ratios (SNRs) are available (Hillebrand and Barnes, 2002). In current cryogen-based SQUID (Superconducting Quantum Interference Device) systems the SNR and spatial resolution is ultimately constrained by the distance between the scalp and sensors that is required for thermal isolation. As these fixed-helmet systems are typically designed for adults, SNR and spatial resolution are further decreased when recording in children. In addition, ictal recordings with SQUID-based helmet systems are problematic due to movement artefacts, and because long-term observations are not feasible. The ability to record seizure activity is of clinical importance, as this provides the most reliable information with regards to the location of the EZ.

Newly developed, cryogen-free, MEG sensors - Optically Pumped Magnetometers (OPMs) - provide an unprecedented opportunity for epilepsy, since they enable non-invasive, wearable recordings with whole-head sensitivity for ictal and interictal activity. OPMs are small and lightweight, yet have a sensitivity that is comparable to that of SQUIDS (Osborne et al., 2018). Importantly, they can be placed directly on the scalp (Colombo et al., 2016; Sander et al., 2012; Shah and Wakai, 2013), thereby improving SNRs (Boto et al., 2016; Boto et al., 2017; Iivanainen et al., 2017) and consequently the sensitivity to, and localisation accuracy of, epileptiform activity. Moreover, wearable OPM-based arrays could allow recordings to be made during the ictal period, and also open up the possibility for long-term observations (Alem et al., 2014).

OPM sensors are passive magnetic field sensors: the transmission of laser light through a gas cell containing a vapour of spin-polarised rubidium atoms reduces in the presence of an external magnetic field, thus providing a highly sensitive measure of the local magnetic field (Tierney et al., 2019). The technique of using alkali vapour cell OPMs to measure magnetic fields is over five decades old (Dupont-Roc et al., 1969), yet in the last decade, since the innovation of high performance semiconductor lasers and miniaturisation of optics, OPMs have been miniaturised and commercialised to a footprint and performance suitable for MEG. The viability of the OPM technology in healthy human subjects has recently been demonstrated (Boto et al., 2018), and this breakthrough work has been followed-up by an increasing body of work with OPMs (Brookes et al., 2022; Tierney et al., 2019), including, for example, their use in children (Hill et al., 2019), assessment of sensory and motor modalities (Borna et al., 2020; Boto et al., 2019; Iivanainen et al., 2020; Roberts et al., 2019), language lateralization and localisation (Tierney et al., 2018), speech processing (de Lange et al., 2021), and the estimation of functional interactions between brain regions (Boto et al., 2021). The potential advantages of OPMs in a clinical setting have also been recognized, with several groups demonstrating their utility in epilepsy. Alem and colleagues used OPMs to record IEDs in a rat model of epilepsy (Alem et al., 2014), with conformation from intracranial electrical recordings. Feasibility in humans has been demonstrated with a single adult patient (Vivekananda et al., 2020). However, ictal recordings and a direct comparison with SQUID-MEG recordings were not performed. More recently, Feys and colleagues reported on the relative merits of OPMs in childhood epilepsy, showing that compared to SQUIDs the IEDs recorded with OPMs had higher SNR for 4 out of the 5 children studied, and higher amplitude for all (Feys et al., 2022).

The theoretical advantages of OPMs over SQUID-based MEG, which include higher SNRs and more accurate source reconstructions, have been demonstrated in modelling (Boto et al., 2016; Iivanainen et al., 2017) and experimental (Boto et al., 2017; Feys et al., 2022) studies. To demonstrate the advantages of higher SNRs in a clinical setting we studied six patients with focal, drug-resistant epilepsy, with previously characterised sources of IEDs: three adults with deep or weak sources, and three children with focal epilepsy, in order to demonstrate that by using on-scalp sensors instead of the standard, adult-sized, helmet the sensitivity to, and SNR of, epileptiform activity increases; a seventh, adult patient with frequent seizures was included in order to demonstrate another advantage of a wearable MEG system, namely that it enables seizure-recordings.

## 2. Methods

### 2.1 Patients

We included seven patients with focal, drug-resistant epilepsy who had already undergone a successful clinical SQUID-based MEG at the Amsterdam UMC, location Vrije Universiteit Amsterdam, as part of their clinical workup for epilepsy surgery. The clinical SQUID-based MEG was deemed successful if the patient did not have claustrophobic or anxiety experiences, was cooperative and not restless, and did not cause many artefacts due to e.g. orthodontic material, and if IEDs could be identified. Three children 10-12 years of age were included. The four adult patients had also undergone invasive EEG recordings (stereo-EEG; sEEG), that were used to confirm the irritative zone as identified with MEG. One patient was selected because he had daily seizures. sEEG, in combination with seizure semiology, was used to confirm the seizure onset zone for this patient. None of these patients had undergone surgery for their epilepsy, because the hypothesised EZ was either bilateral in the mesial temporal lobes, multifocal, or near somatosensory areas, or for socioeconomic reasons. Patients used their regular anti-seizure medication on the day of the recordings, and were sleep deprived in order to increase the incidence of IEDs (Malow, 2004) (see Table 1 for details). Written informed consent was obtained from patients and/or their caretakers at inclusion, and the study was performed in accordance with the Declaration of Helsinki and approved by the VUmc Medical Ethics Committee.

**Table 1:**
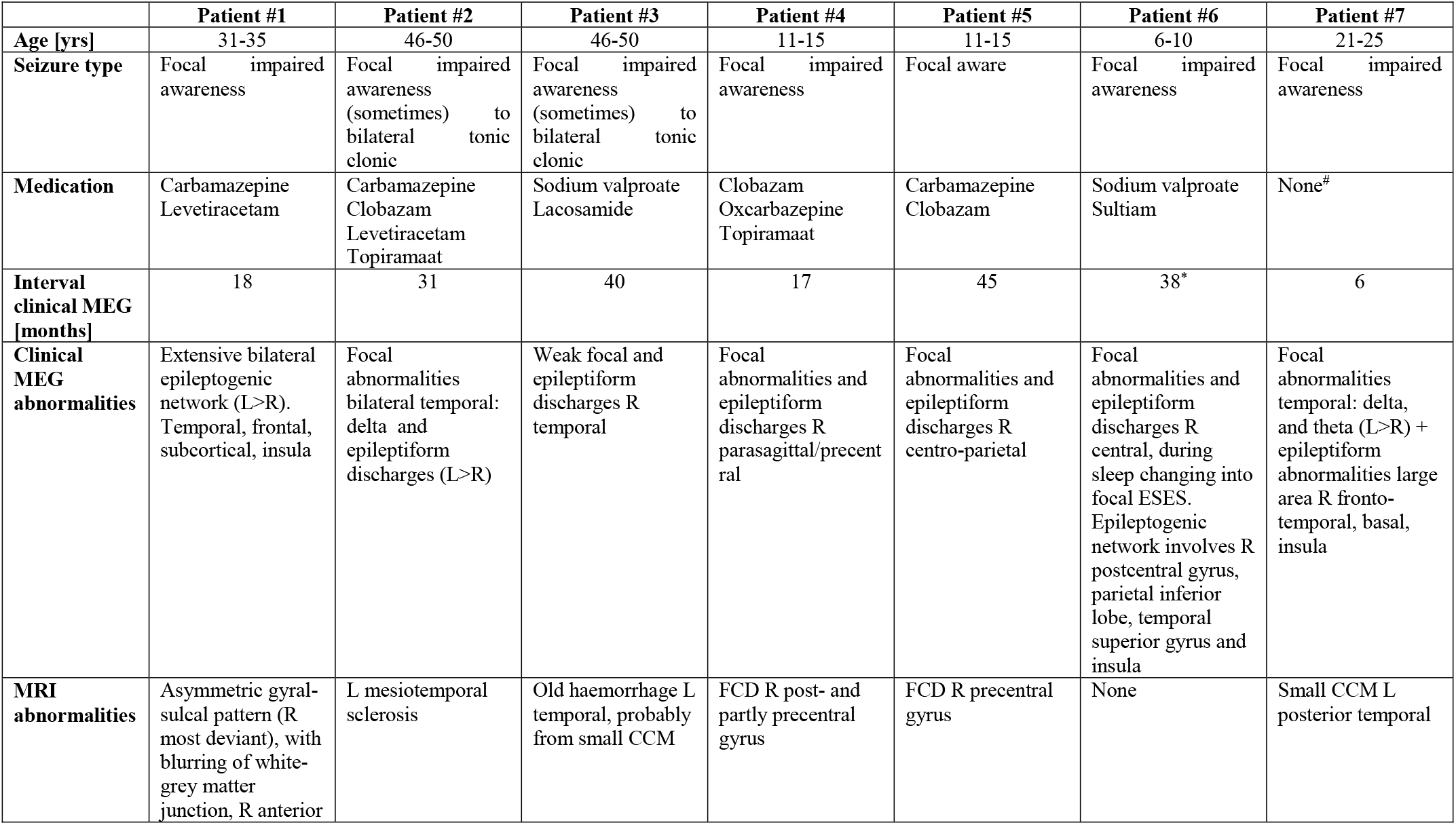
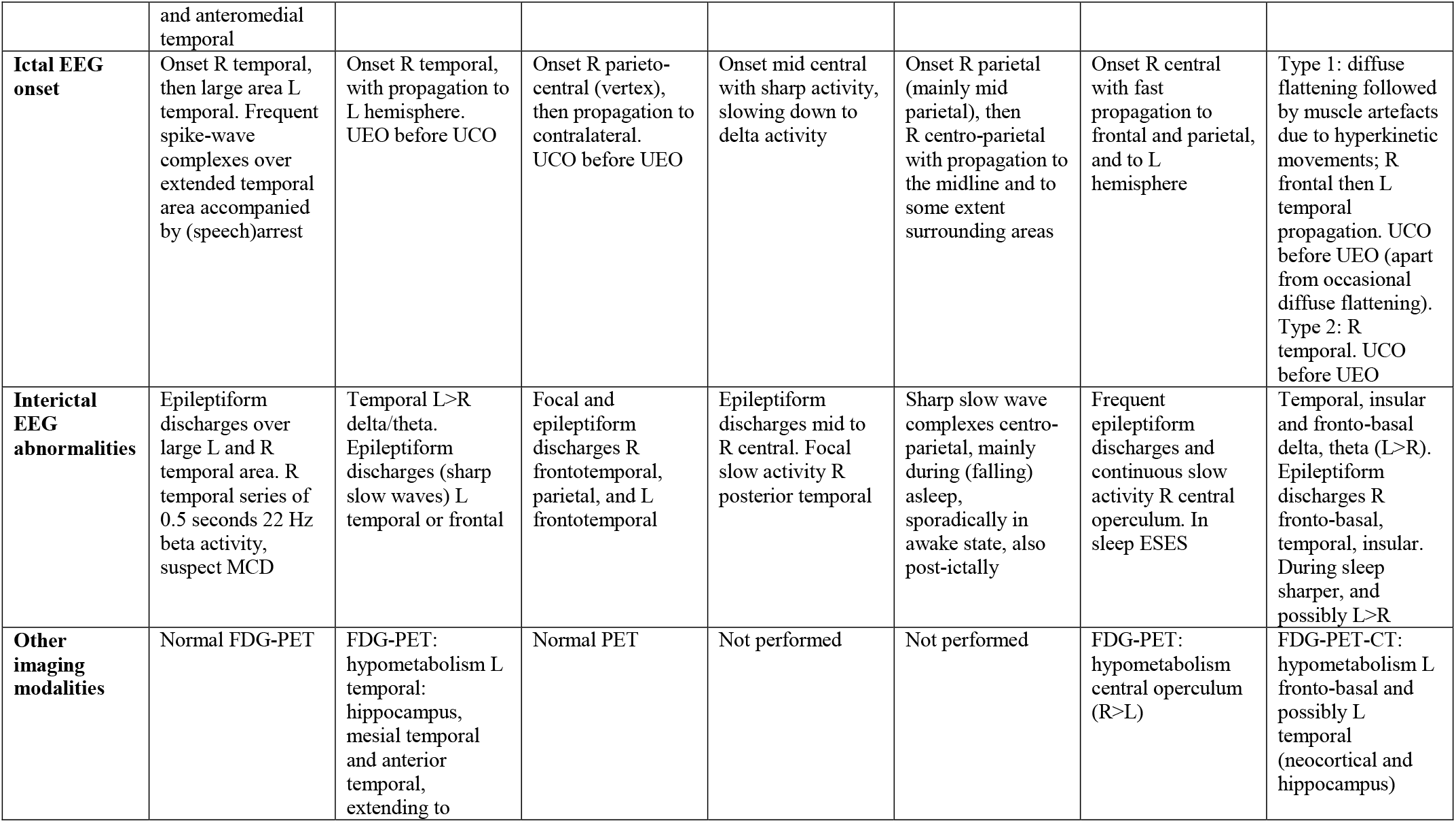
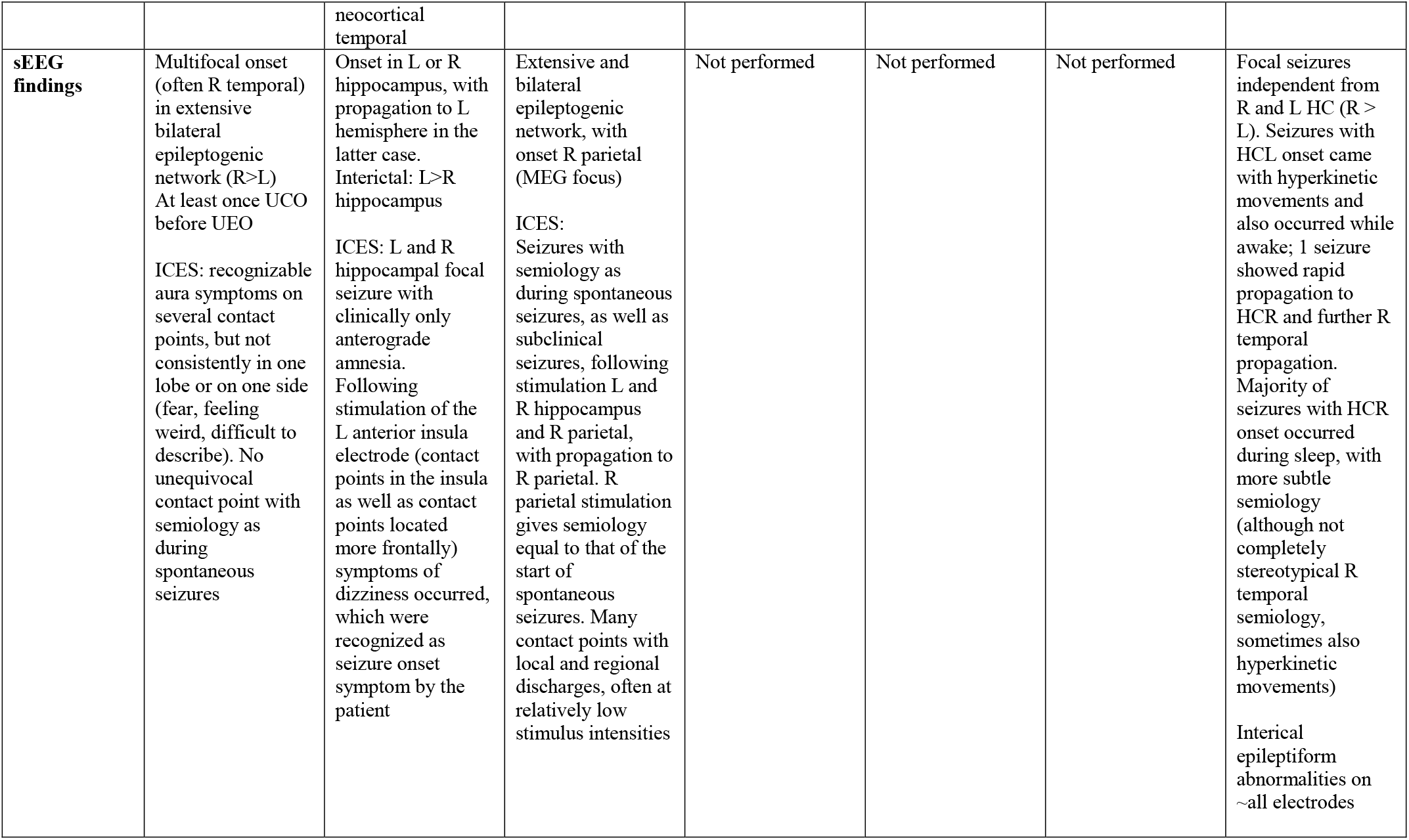
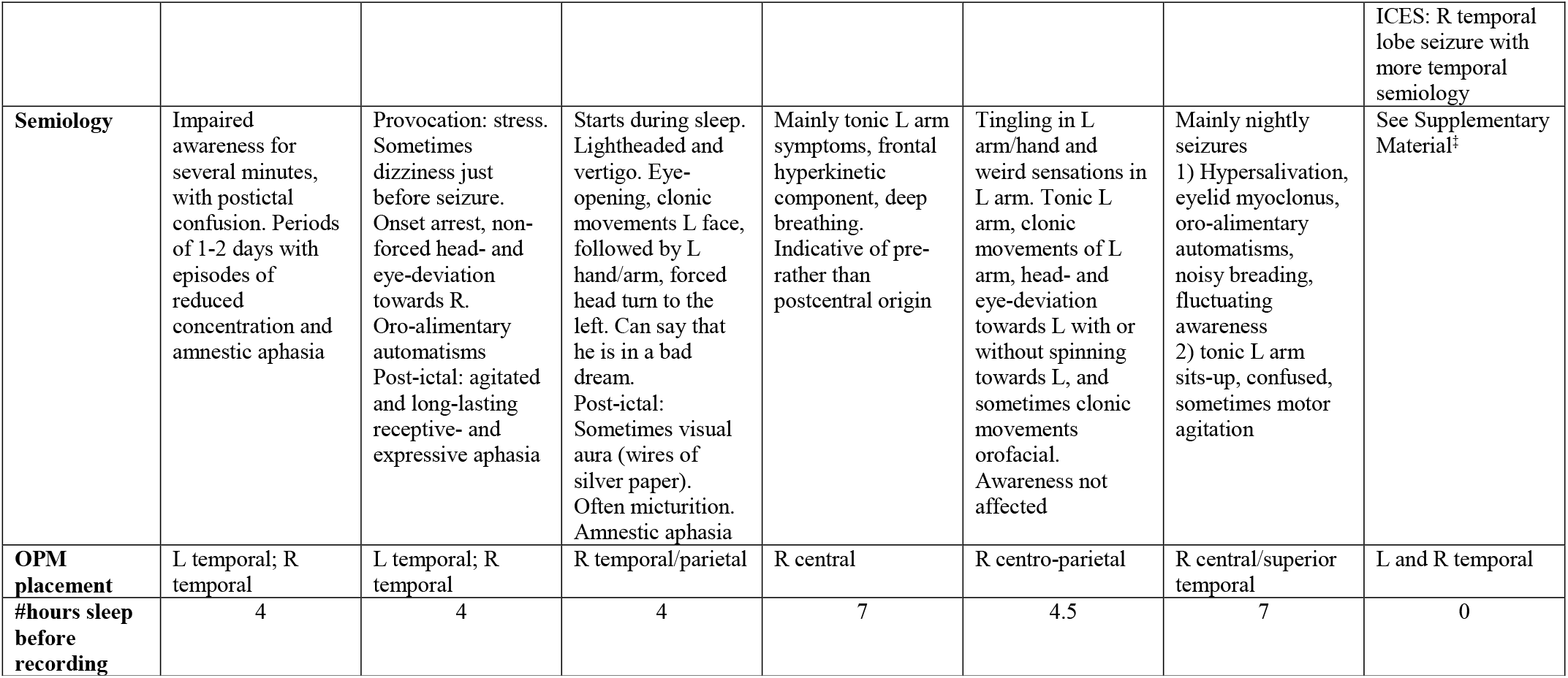
Patient characteristics and recording preparation, showing the location of IEDs in MEG and EEG, ictal EEG onset, interictal and ictal sEEG findings, PET and CT abnormalities, MRI findings, and semiology. All patients were male, and all patients slept less than usual the night before the recordings. OPM placement indicates where the OPMs were placed. L = left; R = right; CCM = Cerebral Cavernous Malformation; CT = Computed Tomography; ESES = Electrical Status Epilepticus during slow-wave Sleep; FDG-PET = Fluorodeoxyglucose-Positron Emission Tomography; HC = hippocampus; ICES = IntraCranial Electrical Stimulation; MCD = Malformation of Cortical 181 Development; MRI = Magnetic Resonance Image; REM = rapid eye movement; UCO = unequivocal clinical onset; UEO = unequivocal electrographic onset; ^*^contained artefacts due to orthodontic material, yet data was interpretable; ^#^after trying many: anti-seizure medication resulted in either one or more of the following: itchiness, eczema, drowsiness, memory problems, mood disturbances, fear, and had no sustained effect on the severity or frequency of seizures; ^‡^on average 8 seizures/night in the weeks before the OPM recording.

### 2.2 OPM setup, recordings and analyses

#### 2.2.1 OPM sensors

Six, commercially available Gen2.0 OPMs (QuSpin Inc, Louisville, CO, USA; Zero field magnetometer, 2^nd^ Generation, dual-axis measurement), with sensitivities of 7-13 fT/√Hz and a dynamic range of ±5 nT (Osborne et al., 2018), were used. We recorded simultaneously along both the radial and a tangential axis to increase the number of measurements and to increase the separability of neuronal and noise signals (Brookes et al., 2021; Nurminen et al., 2013; Rea et al., 2022; Tierney et al., 2021a; Tierney et al., 2022), at a cost of a slight reduction in sensitivity (Osborne et al., 2018). The sensors operate in the spin exchange relaxation-free regime, and the (near) zero-field environment is achieved through ‘on-sensor’ electromagnetic coils wrapped around the vapour cell, that can compensate for remnant fields in the magnetically shielded room (MSR) of up to 50 nT. Before a recording was started, these coils were activated and optimised for a fixed sensor position and orientation, using QuSpin’s QZFM UI acquisition software (version 6.5.10; using [X, Y, Z]-field zeroing and analog output gain of 0.33). Data were recorded with a sampling frequency of 600 Hz using a National Instruments 16-bit NI-9205 ADC interfaced with a LabVIEW (National Instruments (NI) Corporation, Austin, TX) programme developed at the University of Nottingham. The same software was also used to control coil-drivers (QuSpin Inc), using a 16-bit NI-9264 DAC module, for dynamic noise compensation (see below).

#### 2.2.2 Noise compensation

All MEG recordings were performed in a MSR (Vacuumschmelze GmbH, Hanau, Germany) that houses both the OPM-system and a 306-channel cryogenic system (Triux Neo; MEGIN OY, Espoo, Finland). The cold-head, which is part of the internal helium liquefier of the cryogenic system, causes static fields with a magnitude of ∼300 nT, which is outside the OPMs operational range of 50 nT. We therefore installed a set of coils around the cold-head to minimise the field and gradients in the direction of the long-wall of the MSR (Figure 1). The magnitude of the remnant fields in the MSR was reduced to ∼30 nT, using a maximum current of 4A (to avoid overheating) from a low-noise power supply (HMP2020; Rohde & Schwarz GmbH & Co. KG, Munich), fed through an RC lowpass filter unit (-3dB at 0.2 Hz) to reduce current noise. A set of 5 bi-planar coils (Holmes et al., 2019) was then used to bring the remnant fields within the dynamic range of the OPMs (Figure S1). This coil set generates the three uniform field components and five (linear) magnetic field gradient components to produce a magnetic field which is equal and opposite to that experienced by the OPM array. To compensate the field, two three-axis fluxgates (Bartington Instruments Ltd, Witney, UK; MAG-13MSQ100) were used as reference sensors, and placed at two diagonally opposite corners of the virtual nulling-volume (Figure 1). The output of the fluxgates was digitised using a 24-bit NI-9207 ADC module and visualised using the LabVIEW programme. Through manual adjustment of the coil-drivers the remnant fields could be reduced to ∼1 nT, following which the fluxgates were removed and OPM recordings could be performed. Due to fluctuation in our inner-city environment, field levels would typically reach levels of ∼4 nT again within a few minutes (Figure 2).

**Figure 1:**
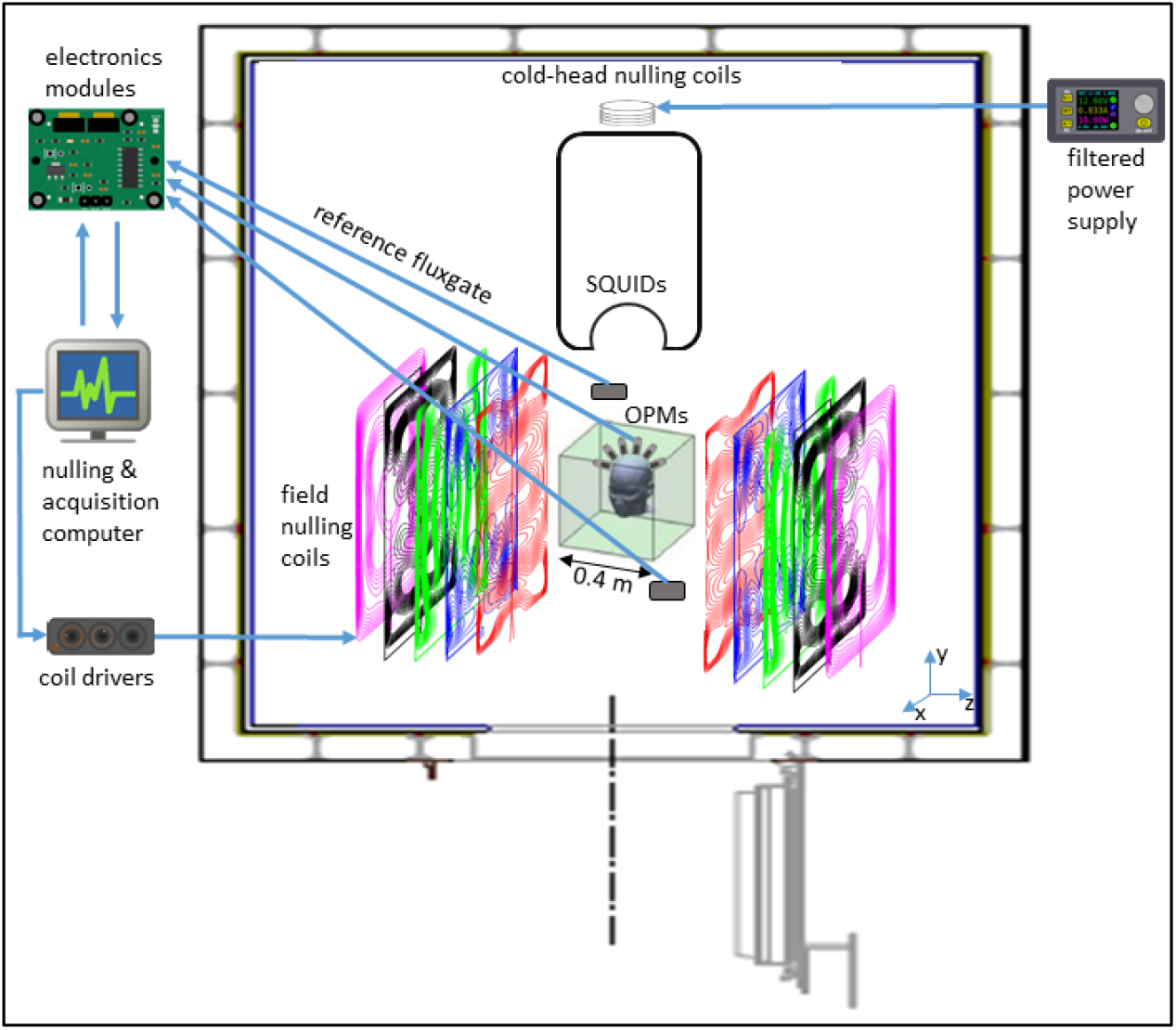
System setup. The whole OPM-system was placed in a magnetically shielded room, together with the SQUID-based system. The cold-head, which is part of the helium liquefier of the SQUID-system, caused static fields with a magnitude of ∼300 nT. Compensation coils around the cold-head reduced these fields to ∼30 nT. Field-nulling coils were wound on five large planes placed either side of the participant, different coloured wirepaths show coils designed to produce different field components (shown deliberately offset here; see also Figure S1). Two fluxgates, placed near the location of where the patient’s head will be during the recordings, were used to record the remnant (static) background fields, and the user manually adjusted the current through the field-nulling coils in order to bring the remnant field level down to ∼1 nT. During the patient-recordings, the low-pass (<3 Hz) filtered signals from the OPMs themselves were used to dynamically compensate for temporal variations in the remnant fields, so that the field experienced by the OPMs in a typical recording remained below 0.4 nT.

**Figure 2:**
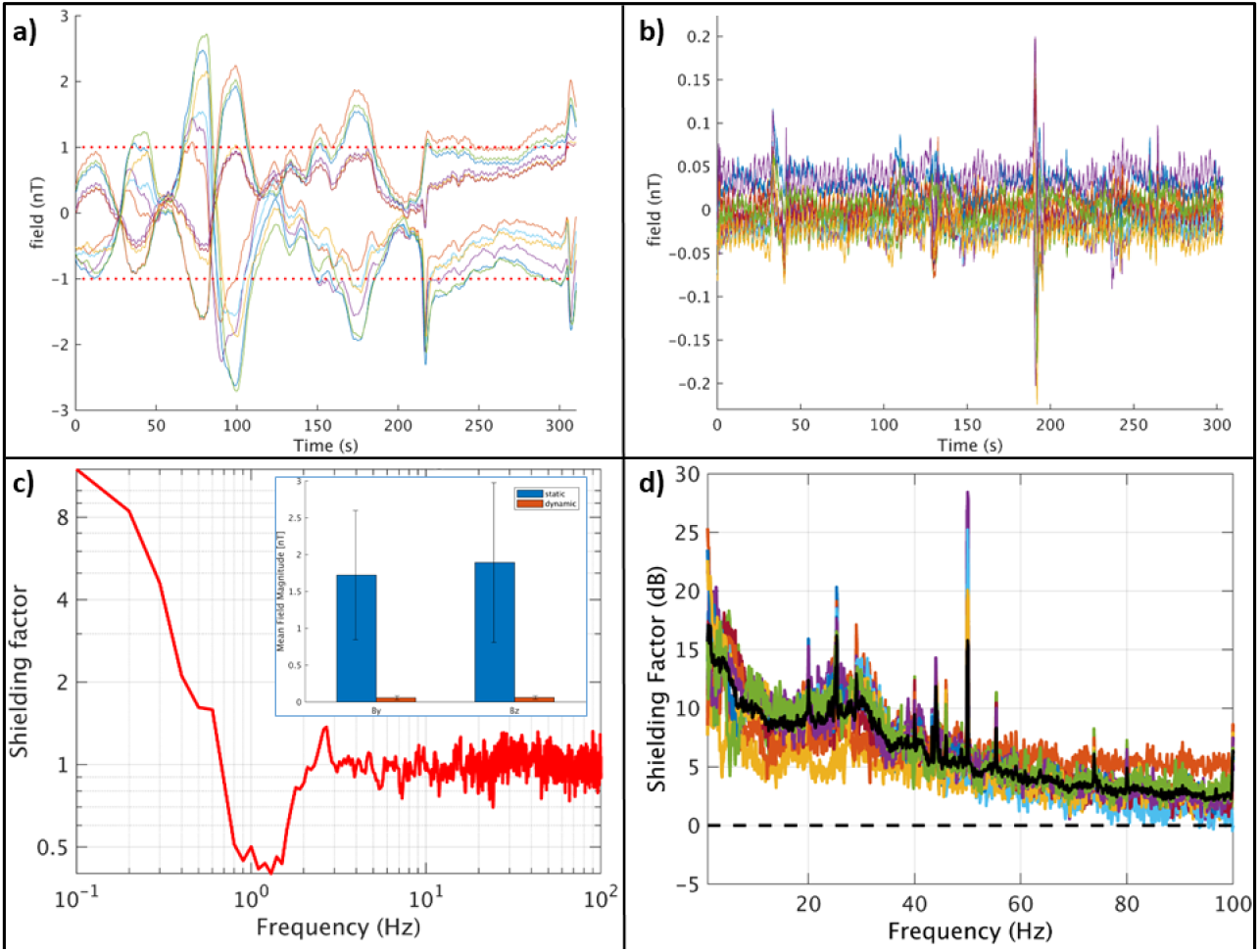
Performance of static and dynamic nulling, and Homogenous Field Correction. Before the patient recordings, the dynamic nulling performance was quantified with a 5-minute empty-room recording with the OPMs in the patient-helmet (here: patient #5). Panel **a** shows the data for all 12 channels with only compensation of the static remnant magnetic field (using internal and external coils). Note that the remnant fields did not remain below 1 nT throughout the recording due to fluctuations in the environmental magnetic fields. However, when dynamic nulling was applied (panel **b**), the change in field could be kept below 0.3 nT. The shielding factor (panel **c**; computed as the power spectral density for the dynamic nulling divided by the power spectral density for the static nulling) was above 1 for frequencies below 0.7 Hz, with a maximum of 12 for 0.1 Hz, and approximately 1 above 2.5 Hz. In between 0.7 and 2.5 Hz the shielding factor was smaller than 1, which is due to noise that is introduced by the choice of the PID-controller’s gains. The inset shows the field magnitude (L2-norm) of the field for the 12 channels (By- and Bz-direction separately) averaged over time (with error-bars showing the standard deviation) with static (blue) and dynamic (red) nulling applied, showing that dynamic nulling decreased the field magnitude with a factor 30. The field magnitude averaged over the empty-room recordings for the 7 patients was 0.09 and 0.11 nT for By and Bz, respectively (not shown), with the maximum absolute field in a channel never exceeding 0.7 nT. Panel **d** shows how Homogenous Field Correction further removes noise from the recorded data (recording 1 from patient #5). The black line denotes the HFC shielding factor (in dB) averaged over all channels (coloured lines).

Movement of the sensors through such remnant fields during a patient recording would send them outside their dynamic range, and even without such movements the remnant fields would degrade the signal fidelity through cross-axis projection errors (Borna et al., 2022). We therefore used dynamic compensation (Holmes et al., 2019; Iivanainen et al., 2019), based on the OPMs themselves, to reduce the remnant fields further and to keep them stable during an experiment. First, the response of the OPMs to a known current was determined by sequentially sending pulses (50 msec, 0.4 V) to the coils, resulting in an 8 (coils) x 12 (sensors) calibration matrix. During a recording, the inverted calibration matrix was used to set the required voltage outputs for the coil-drivers to minimise the sum-of-squares of the sensor outputs. A proportional–integral– derivative (PID) controller was used to drive the sensor outputs towards zero, using manually tuned proportional and integral gains (derivative gains were set to zero). As input to the PID, the averages of 20-sample segments of data for all 12 channels were used, sampled at 600 Hz and digitally filtered at 3 Hz with a fifth-order low-pass Butterworth filter (chosen so that the controller would not remove the (brain) signals of interest from the OPM recordings). This reduced the maximum field changes experienced by the sensors to <0.4 nT (Figure 2) during empty-room recordings. During patient-recordings, the reduced static remnant fields and gradients ensured that head-movements were better tolerated (Boto et al., 2018; Holmes et al., 2018), and the dynamic compensation ensured that remnant fields/gradients remained small throughout the recordings.

#### 2.2.3 3D-printed helmets

In order to keep the sensors firmly in place and on the scalp, individualised rigid 3D-printed helmets were constructed on the basis of the patients’ anatomical magnetic resonance images (MRIs) that were available from the clinical workup. These were typically T1-weighted images recorded using a 3T scanner, with ∼1 mm resolution. The scalp-surface was extracted from these MRIs, and a standard helmet design was projected onto this surface (for the children a 2mm offset was added in order to account for growth during the period (2-4 years) between MRI- and OPM-scan) (Siemens NX, version 1953; Siemens AG) in order to create a patient-specific helmet-model. The helmet-model contained a removable cap at the front so that the helmet could slide easily over the head, and openings for a chin strap to enable firm fixation of the helmet on the head (Figure 3). Individual OPM-holders, which included flexible side-legs to ease removal of the OPMs, were manually added to the helmet-model such that the OPMs would sample the dipolar field patterns that were produced by IEDs that were identified in the previously recorded clinical MEG. For the adult patients, the seizure onset zone, as identified in the stereo-EEG recordings, was also used to guide placement of the OPM-holders. The location of the vapour cell within the OPM casing was accounted for, assuming that OPMs were placed within the holder such that one sensitive axis was perpendicular to the head, and the other parallel to the head in the nasion-inion direction. Three reference points were added to the model to enable co-registration of the helmet with the patient’s head. The helmet-models were subsequently 3D-printed using biocompatible sintered PA12 (polyamide) nylon (Oceanz, Ede, The Netherlands). Before the patient recordings, the crosstalk between the OPMs was determined for each helmet design by sequentially activating the on-board coils and recording the responses of the remaining OPMs (see (Boto et al., 2018) for details). The maximum crosstalk for the different helmets was on average 1.4% (range: 0.90 - 1.91%), and was therefore not considered during further analyses (Boto et al., 2018; Tierney et al., 2019).

**Figure 3:**
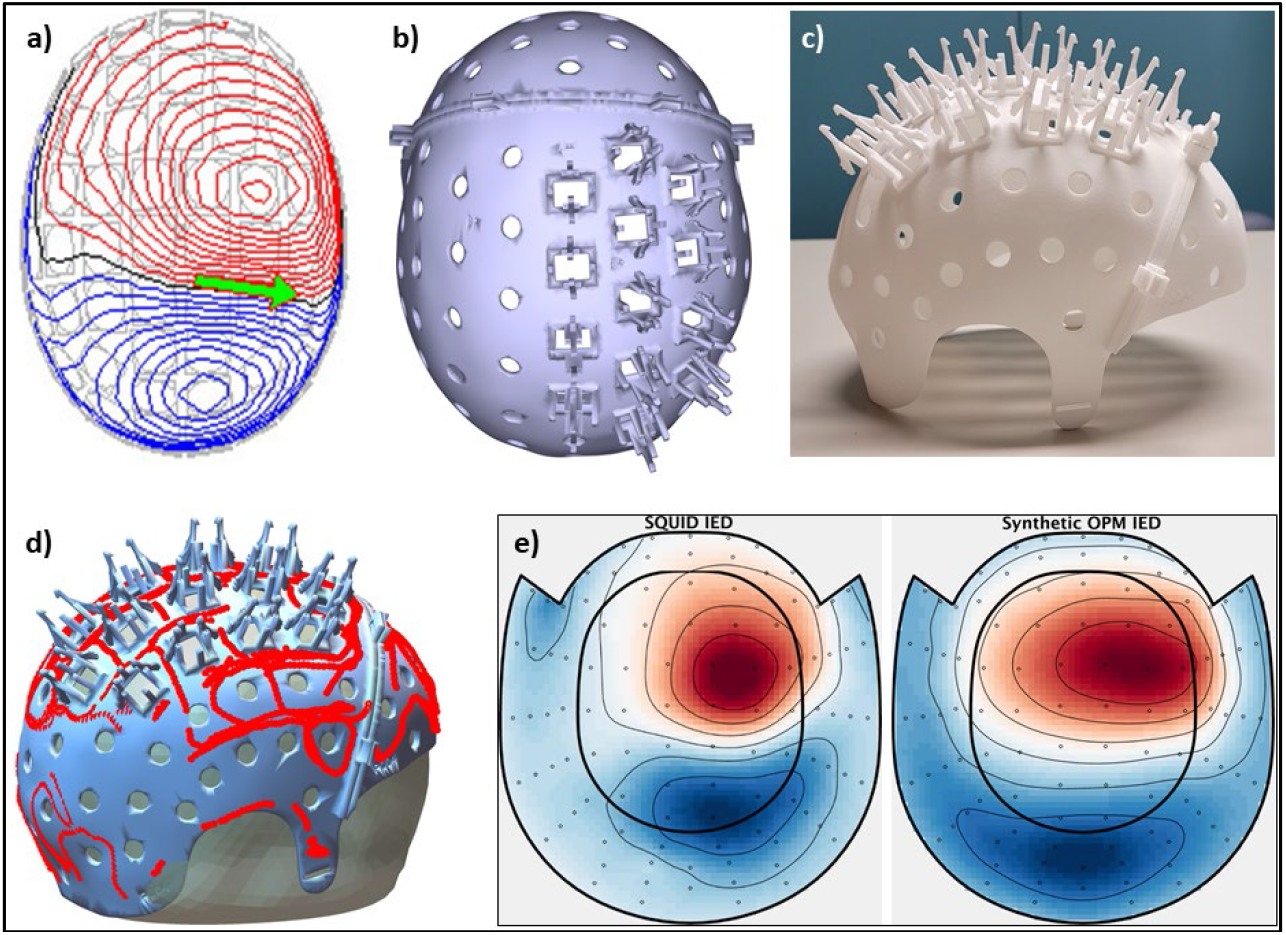
Helmet design for patient #4 and field patterns recorded with SQUIDS and OPMs. **a)** Field pattern produced by IED (green arrow indicates the source-reconstructed equivalent current dipole) in the previously recorded clinical MEG, originating from, and in agreement with, a right central focal cortical dysplasia. **b)** 3D-helmet model, including removable front and OPM-holders. **c)** 3D-printed helmet. **d)** Digitised helmet points aligned with helmet-model, and co-registered to the anatomy (head surface from MRI). e) Field pattern for an IED recorded with the SQUID-based system (left), as well as field pattern for an IED recorded with the OPMs, projected onto the SQUID-sensor layout (after inverse/forward projection with minimum norm (Knösche, 2002; Marhl et al., 2022)). Note the good agreement between the IED field patterns, despite the limited sampling with the OPMs, suggesting that both systems recorded the same phenomenon. Also note the agreement with the previously recorded IED (panel a).

#### 2.2.4 MEG-MRI co-registration

Knowledge of the positions and orientations of the OPMs with respect to each other and the anatomy of the brain is required for source-reconstruction and array-based post-processing. The positions and orientations of the OPMs with respect to the helmet are known from the helmet modelling (assuming no errors in 3D-printing). The position of the helmet with respect to the head was determined by digitising the reference points on the 3D-printed helmet, as well as the nasion and pre-auriculars, the nose, outline of the helmet, and the forehead (when not fully covered by the helmet), using a 3D digitizer (Fastrak; Polhemus, Colchester, VT, USA). A rigid transformation of the three reference points then aligns the helmet (and OPMs) with the head (Figure 3). The head was co-registered to the patient’s anatomical MRI through surface matching of the head surface as extracted from the anatomical MRI with the digitised nose and forehead, or though matching of the nasion and pre-auricular points when surface matching was not possible. Combining the two transforms provides co-registration of the OPMs with the brain anatomy (Figure 3).

#### 2.2.5 OPM recordings

Recordings were performed in the morning in seated position, with the OPM sensors in the centre of the virtual nulling-volume (Figure 1). Patients were instructed to sit still, with eyes closed, and were allowed to fall asleep during the recording. The calibration matrix for dynamic field compensation was determined, following which a 15-minute OPM recording was started, as well as recording of the video-signal from the patient-monitoring system. The performance of the dynamic field compensation was monitored online, and if the noise compensation became unstable (due to large movements or a new head position that was incompatible with the calibration matrix), meaning that the feedback signal would diverge, a new recording was started. For patients with known bilateral IEDs, separate recordings were performed with OPMs over the left or right hemisphere (Table 1). For patient 7, three sensors were placed over each hemisphere. Because of the known semiology and to avoid the possibility that the patient would hurt himself or damage the OPM cabling, movement was restricted by a belt around the chest and bandages around the wrists. Recordings for this patient lasted until a seizure was recorded.

#### 2.2.6 OPM pre-processing

Despite the dynamic field compensation, the OPM recordings still contain interference from external (and internal) sources, as well as due to movement of the sensors through the remnant fields when the patients move their heads. Sophisticated spatial filtering techniques for noise removal, such as Signal Space Separation (Taulu et al., 2004), could not be applied to the 12 channel OPM-recordings, as the magnetic fields are undersampled and, more fundamentally, because the internal space basis set is invalid as a model of brain signals for on scalp sampling (Tierney et al., 2022; Wens, 2022). Tierney and colleagues have shown that as a first approximation the magnetic interference can be modelled as a spatially homogenous field, which can subsequently be regressed from the data (Hill et al., 2022; Seymour et al., 2022; Tierney et al., 2021a). Despite the low number of OPMs, noise was effectively removed, typically resulting in shielding by 15 dB at low frequencies (<2 Hz), line noise, and for artefacts with a sharp peak in the noise spectrum (24 Hz in our environment), 5-10 dB for frequencies between 2 and 50 Hz, and 5 dB and slowly declining above 50 Hz (Figure 2).

#### 2.2.7 OPM source reconstruction

We applied beamforming (Hillebrand and Barnes, 2005; Hillebrand et al., 2005) because of its ability to remove interference (Adjamian et al., 2009) (Figure S4) and to align with our clinical work-flow (Hillebrand et al., 2016a), yet realising that the beamformer’s ability to localise activity will be limited with a small number of sensors. The DAiSS toolbox in SPM (version 12) was used to reconstruct the time-series of neuronal activity (so-called virtual electrodes) for the centroids (Hillebrand et al., 2016b) of 246 regions of the Brainnetome atlas (BNA) (Fan et al., 2016). Broadband (0.5 – 48 Hz) beamformer weights were constructed, for which the data covariance matrix was filtered using a discrete-cosine-transform after applying a Hanning taper, 5% Tikhonov regularisation was used when inverting the data covariance matrix. The lead fields were based on an equivalent current dipole source model with optimum orientation (Sekihara et al., 2004), and a single shell head model (Nolte, 2003) based on the inner skull-surface of the co-registered MRI, and the homogenous field correction was taken into account (Hipp and Siegel, 2015; Tierney et al., 2021a). Sensor-level data, filtered in the 3 – 48 Hz band with a fifth-order Butterworth filter, were subsequently projected through the normalised beamformer weights (Cheyne et al., 2006).

### 2.3. SQUID setup, recordings, and analyses

#### 2.3.1 SQUID setup

The SQUID-recordings were performed with the cryogenic whole-head system, using our standard clinical protocol for epilepsy (Hillebrand et al., 2013). This includes the recording of two horizontal and one vertical electrooculography channel and an electrocardiography (ECG) channel, and continuous recording of the head position relative to the MEG sensors using signals from five head-localization coils. The positions of the head-localization coils and the outline of the patient’s scalp and nose (∼4000 points) were digitized using the 3D digitizer. These scalp/nose points were used for co-registration with the head surface as extracted from the patient’s anatomical MRI, using surface matching.

#### 2.3.2 SQUID recordings

Recordings were performed in the afternoon in supine position, after switching off the cold-head compensation coils and bi-planar field-nulling coils. Data were recorded with a sample frequency of 1000 Hz, with an anti-aliasing filter of 330 Hz and a high-pass filter of 0.1 Hz. Internal active shielding (IAS) (Taulu et al., 2019), using MEGIN’s in-wall feedback-coils, was used for patient #1, #2, and #5, but unavailable for the other patients due to technical problems. Four datasets of 15-minute duration were recorded in a task-free eyes-closed condition, during which the patients were allowed to fall asleep.

#### 2.3.3 SQUID pre-processing

Cross-validation Signal Space Separation (xSSS) (van Klink et al., 2017) was applied to aid visual inspection of the data. Channels that were malfunctioning, for example due to excessive noise, were identified by visual inspection of the data by A.H. (mean number of excluded channels was 8, range 6–10), and removed before applying the temporal extension of SSS to the raw data (MaxFilter, version 2.2.15; Elekta Neuromag Oy) (Taulu and Simola, 2006), using a subspace correlation limit of 0.9 and a sliding window of 10 sec.

#### 2.3.4 SQUID source reconstruction

Our default, atlas-based beamforming implementation was used (Hillebrand et al., 2012; Hillebrand et al., 2016b). Elekta’s beamformer (version 2.1.28) reconstructed the time-series of neuronal activity for the centroids of the parcels in the BNA atlas. Broadband beamformer weights were computed, for which the data were filtered using a single-pass FIR filter in MaxFilter, using a Kaiser window with an order of 10000 and 104, and attenuation of 60 dB at 0.35 Hz and 72 Hz, for the high pass (0.5 Hz) and low pass filter (48Hz), respectively. Singular value truncation was used when inverting the data covariance matrix to deal with the rank deficiency of the data after SSS, using a truncation limit of 1e^−6^ times the largest singular value. An equivalent current dipole with optimum orientation (Sekihara et al., 2004) was used as source model, and a single sphere, based on the scalp-surface of the co-registered MRI, was used as head model. The broadband data were subsequently projected through the normalised beamformer weights (Cheyne et al., 2006), after which a 3 Hz high-pass filter was applied in MaxFilter (both at the sensor- and source-level) to enable comparison with the OPM data.

### 2.4 IED detection and quantification

IEDs were visually identified at sensor- and source-level and marked by an experienced EEG/MEG technician (N.S.). Subsequently, an automatic algorithm (see Supplementary Material) was used to quantify the SNR of the IEDs at sensor-level, and to identify IEDs that were missed on visual inspection. A second assessor (A.H.) removed false positives from the automatically identified IEDs, using the waveforms and field maps of the visually identified IEDs as references. For the OPM data, field maps of IEDs identified in the SQUID data were used as reference and the OPM data were projected onto the SQUID-sensor layout to ease the comparison and identification of true positive IEDs (Figure 3). The remaining IEDs were compared between SQUID and OPM recordings for each patient in terms of Z-score (averaged over IEDs), as a proxy for SNR, and the spike-wave index (SWI), which is defined here as the percentage of seconds that contained an IED (Aeby et al., 2021). The average Z-scores were compared for each patient using the Mann-Whitney U test and an alpha of .05. Effect size was determined using Cliff’s delta, and categorised as negligible (|d| < 0.147), small (0.147 ≤ |d| < 0.33), medium (0.33 ≤ |d| < 0.474), or large (|d| ≥ 0.474).

## 3. Results

Results of the analysis of the sensor-level OPM- and SQUID-based data are provided in Table 2.

**Table 2:** SNR and frequency of occurrence of epileptiform activity for the OPM- and SQUID-based (gradiometers only) sensor-level data. The SNR averaged (and standard deviation) over IEDs is reported. ^*^based on the datasets that contained epileptiform activity.

|  | OPM |  | SQUID |  | SNR<sub>OPM</sub> vs SNR<sub>SQUID</sub> |  |  |
| --- | --- | --- | --- | --- | --- | --- | --- |
|  |  |  |  |  | Mann-Whitney U | Cliff's d | p-value |
| Patient | SNR | SWI | SNR | SWI |  |  |  |
| #1 | 4.47<br>(0.43) | 6.42 | 4.57<br>(0.46) | 10.25 | -3.81 | -0.14 | 1e-4 |
| #2 | - | 0 | - | 0 | - |  | - |
| #3 | 3.85<br>(0.32) | 9.00* | 3.93<br>(0.35) | 6.76 | -2.86 | -0.14 | .0043 |
| #4 | 4.05<br>(0.48) | 11.03 | 4.39<br>(0.61) | 24.50 | -9.53 | -0.36 | 2e-21 |
| #5 | - | 0 | - | 0 | - |  | - |
| #6 | 6.15<br>(0.98) | 0.53* | - | 0 | - |  | - |

Patient #1 showed many short and long series of spike-wave complexes in both the OPM and SQUID data, over both the left and right temporal lobes (Figure 4). Five recordings were performed with the OPMs over the left hemisphere (average duration 709 sec; range: 306-906 sec), and four with the OPMs over the right hemisphere (average duration 811 sec; range: 716-901 sec). The SWI was higher for the SQUID data (10.25) than for the OPM data (6.42). This could be explained by the presence of (partly) independent left- and right-temporal IEDs in combination with the unilateral coverage during the OPM recordings: when considering only unilateral temporal channels in the SQUID data, the SWI dropped down to 6.11 for the left hemisphere, and 6.58 for the right hemisphere, and thereby became comparable to the SWI of the OPM data. The difference between the SNR of the IEDs in the OPM data and SQUID data was negligible (4.47 versus 4.57; U = -3.81, d = -0.14, *p =* 1e-4).

**Figure 4:**
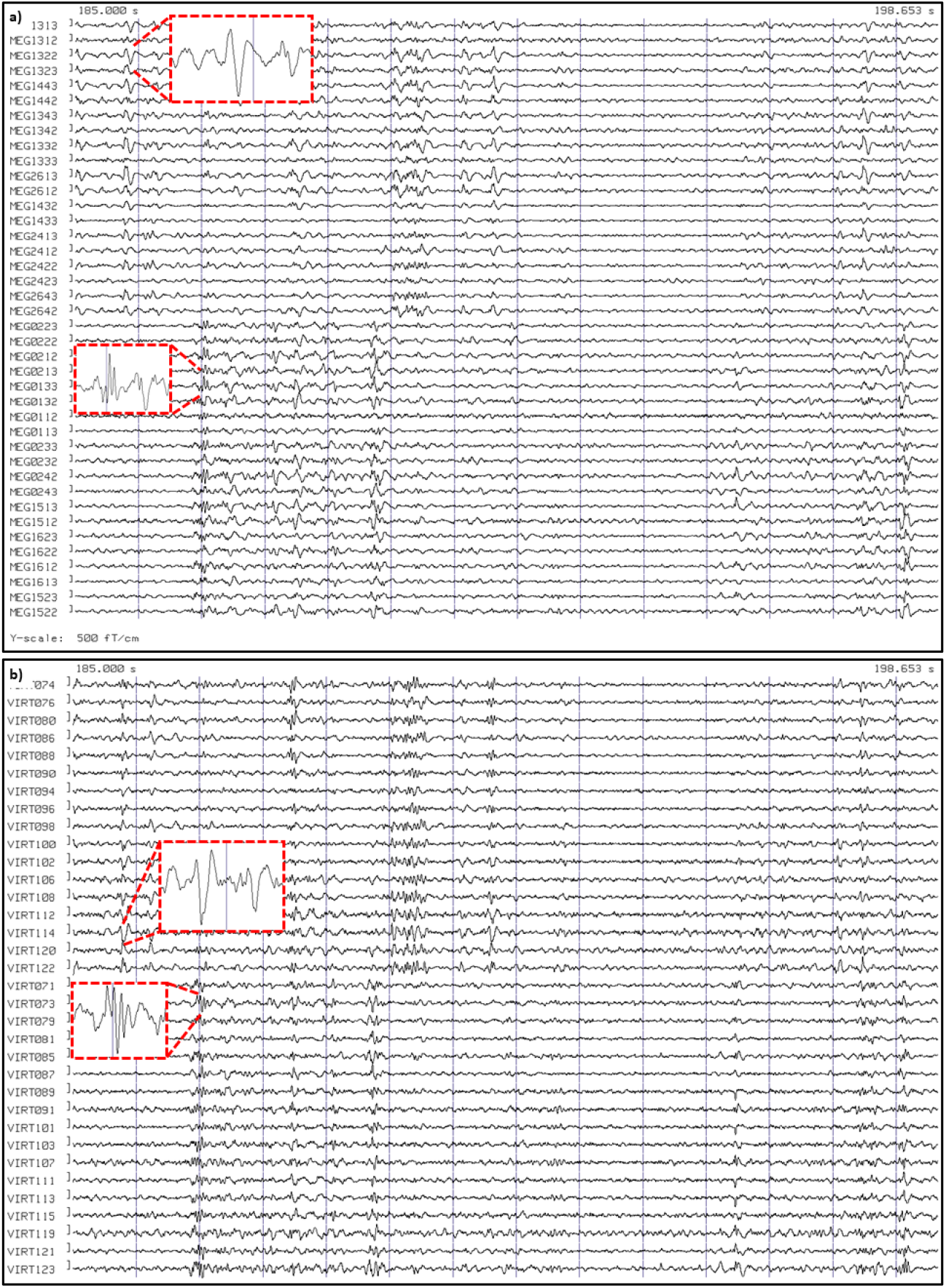
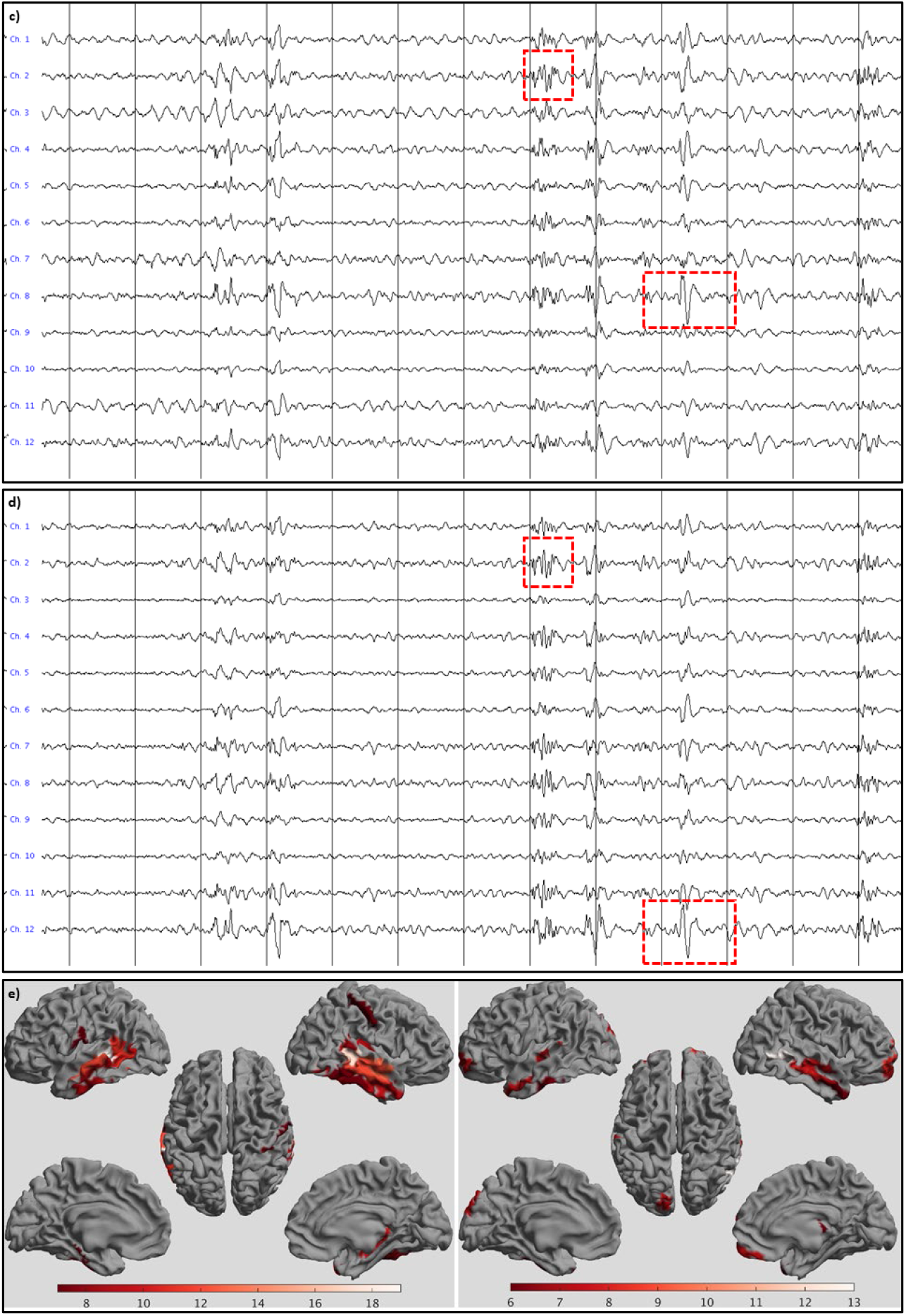
Examples of epileptiform activity for patient #1. IEDs were recorded with SQUIDS (**a, b**) and OPMs (**c, d**) at sensor-level (**a, c**) and source-level (**b, d, e**). **a)** 13.653 seconds of data for a selection of gradiometers over the left (upper half) and right (bottom half) temporal lobes. The grey vertical lines mark 1 second of data, filtered between 3 - 48 Hz. Note the presence of (many) IEDs over both hemispheres, with some examples highlighted. **b)** Virtual electrodes for a selection of the left (upper half) and right (lower half) temporal ROIs of the BNA atlas. **c)** Comparable signals were recorded with the six OPMs, placed over the left temporal lobe in this case (recorded earlier in the day). Alternating channels show recording in the OPMs’ By and Bz direction. As for the SQUID data, some of the spike-waves and polyspikes are highlighted. **d)** Virtual electrode data for the same data segment (selection of left temporal BNA ROIs). **e)** Number of times a region showed the maximum SNR (over all 246 ROIs) for the events that had been identified at sensor-level (total over all datasets) for SQUID (left) and OPM data (right). Results are displayed, with an arbitrary threshold, as a color-coded map on the parcellated template brain, viewed from, in clockwise order, the left, top, right, right midline, and left midline. Note that for both systems the regions in the temporal lobes most frequently had the maximum SNR for the identified IEDs, consistent with SEEG, EEG, and earlier clinical MEG findings (see Table 1).

For patient #2 no IEDs were identified in either the OPM (four 15-minute recordings with OPMs over the left or right hemisphere) or SQUID data.

The SQUID recordings for patient #3 showed small IEDs, with an average SNR of 3.93 ± 0.35, over the right superior temporal/parietal lobe, often occurring in brief series (SWI = 6.76). Similar IEDs were visible on a single channel in three OPM datasets (average duration 907 sec, range 900-916 sec; SWI = 9.00; average SNR 3.85 ± 0.32). For the 4^th^ OPM-recording, this OPM had been moved to a new position (Figure S2), and IEDs could not be identified in this channel anymore. The difference between the SNR of the IEDs in the OPM data and SQUID data was negligible (3.85 versus 3.93; U = -2.86, d = -0.14, *p =* .0043).

For patient #4, six OPM-recordings, with an average duration of 348 sec (range: 194-614 sec), were performed with the OPMs over the right sensorimotor cortex. 282 IEDs were identified (SWI = 11.03). The four SQUID-recordings revealed more IEDs (SWI = 24.50), with significantly higher SNR and medium effect size (4.39 versus 4.05; U = 9.53, d = -0.36, *p =* 2e-21). Figure 3 shows the field configuration for IEDs recorded with both systems, illustrating that the topography is comparable across systems.

For patient #5, no IEDs were identified in either the OPM (7 recordings; on average 465 sec; range 307-631 sec) or SQUID data.

For patient #6, six OPM-recordings were performed with the OPMs over the right central areas/superior temporal lobe. Two datasets, with a duration of 900 and 609 sec, contained 3 and 5 IEDs, respectively, with an average SNR of 6.15. The other datasets (601, 540, 884, and 173 sec in duration) did not contain IEDs, nor did the four 15-min SQUID recordings. As was the case in the previously recorded clinical MEG, the SQUID data contained artefacts in right-temporal channels due to orthodontic material in the right side of the mouth, but these artefacts were largely removed by tSSS. In the OPM data, artefacts due to movement alone and those due to the orthodontic material were difficult to discern, but these were largely reduced by HFC.

After 2.5 hours, patient #7 had a seizure that was recorded with the OPM-system. The semiology was indicative of an onset in the left temporal lobe, or with a right temporal onset with rapid propagation to the left temporal lobe (see Table 1 and Supplementary Material). The seizure onset was visible in OPM sensors over both the left and right anterior temporal lobe, with no identifiable delay between the two hemispheres (Figure 5). At source-level, seizure activity was visible in both hemispheres (Figure S3). Despite the large amplitude movements of the patient during the seizure, approximately 30-40 cm, the OPMs stayed within their dynamic range (not shown), albeit with large movement-artefacts. Interictally, independent IEDs were observed over left and right temporal lobes, as well as simultaneously over both temporal lobes (Figure S4), consistent with the findings from earlier sEEG recordings and seizure semiology, which pointed at independent SOZs in the left and right temporal lobe, as well as occasional rapid propagation of ictal activity between these regions. Of note, in the previously recorded SQUID-MEG IEDs were found in right fronto-temporal regions, including frontobasal and insula, but no IEDs were found in the left hemisphere. SQUID recordings were not performed during the current visit.

**Figure 5:**
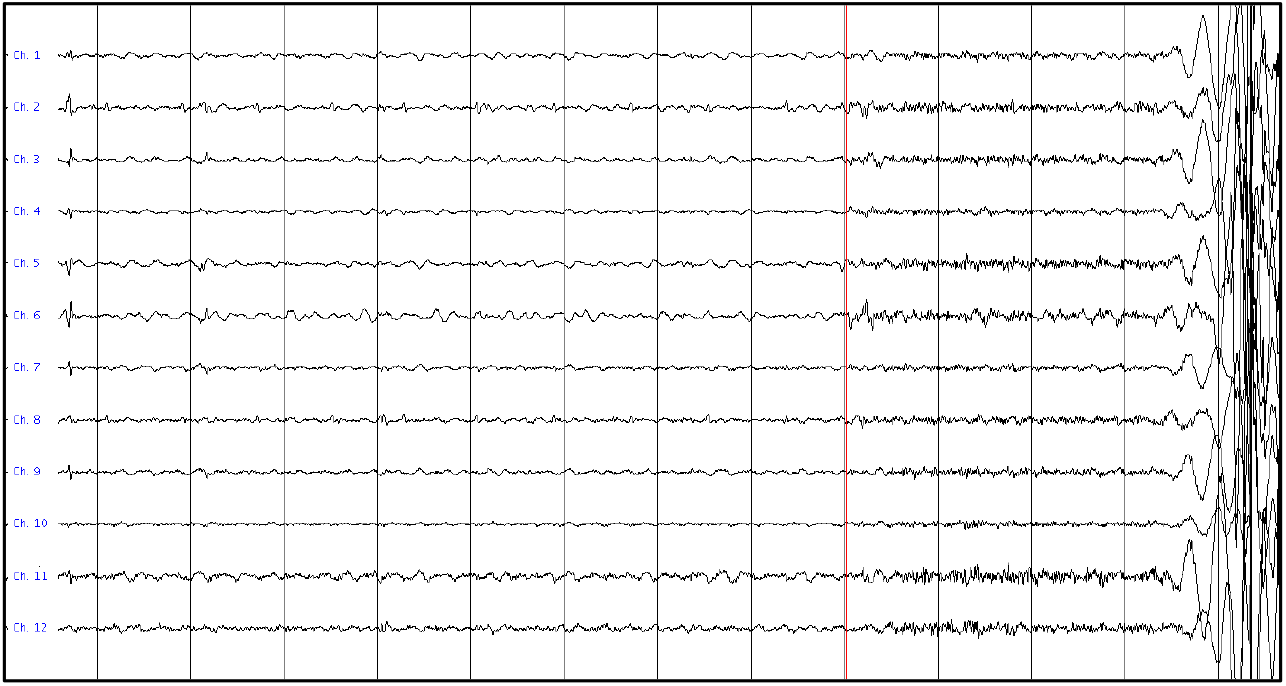
Ictal onset for patient #7. The first and last 6 channels are from the 3 OPMs over the right and left anterior temporal lobe, respectively, with alternating channels recording in the OPMs’ By and Bz direction (which is, in this case, in the anterior-posterior direction and approximately perpendicular to the scalp (inwards), respectively). The grey vertical lines mark 1 second of data, that were filtered between 3 - 48 Hz. Note the increase in fast activity, simultaneously over both hemispheres, after about 9 secs, marking the start of the seizure (red vertical line). This is followed by artefacts due to movement during the seizure. Although it cannot be ruled-out that the fast activity during the 3 secs before the bodily movements was due to muscle activity, we believe that this is unlikely as the clinical onset of the seizure (blinking) started ∼2 seconds after the onset of the fast activity (see Supplementary Material), and no other movements were discernible during that period (compare also with video-EEG recording in Supplementary Material).

Based on the semiology of the recorded seizure and the striking resemblance with activity for seizures of Type 1 as previously recorded using video-EEG and sEEG (Figure S3), the OPM data was consistent with a left temporal ictal onset and propagation, or right temporal ictal onset with rapid propagation to the left temporal lobe and further left temporal propagation.

## 4. Discussion

The main aim of this study was to demonstrate the feasibility of OPM-based MEG in a clinical population. We showed that interictal epileptiform activity can be successfully recorded from both adults and children with epilepsy, and that seizure activity can also be captured. To demonstrate the advantages of a reduced source-sensor distance with OPMs we quantified and compared the SNR and IED yield with a conventional SQUID-based whole-helmet system. In one patient the OPMs detected IEDs that were not found with the SQUID-system. In one patient the spike yield was higher for the OPM data, with negligible difference in SNR compared to the SQUID data. This was also the case for a patient with a spike yield that was comparable to that for the SQUID data (after accounting for unilateral coverage with the OPMs). For one patient the spike yield and SNR were both lower for the OPMs. In two patients no IEDs were found with either system. Importantly, the wearability of OPMs enabled the recording of seizure activity in a patient with hyperkinetic movements during the seizure. The observed ictal onset and semiology was in agreement with previous video- and stereo-EEG recordings. The ability to record seizures non-invasively, with high spatial resolution, is of clinical importance, as ictal activity often provides more accurate information about the epileptogenic zone than interictal activity (Stefan and Rampp, 2020). A more accurate delineation of the epileptogenic zone may improve surgical planning and ultimately lead to improved seizure outcome in patients with refractory epilepsy.

### Comparison between systems

The noise level of OPMs (7-13 fT/√Hz in our case) is higher than for SQUIDS (∼3 fT/√Hz), yet this is compensated for by the increase in signal amplitude due to the on-scalp placement of OPMs (reducing the source-sensor distance by ∼2 cm compared to fixed-helmet SQUID-based systems). Simulations (Boto et al., 2016; Iivanainen et al., 2017) and experimental data (Boto et al., 2017; Feys et al., 2022) have shown that the SNR of OPM-based measurements is indeed higher than for SQUID-based recordings. The largest gains are found for cortical sources, as the relative reduction in source-sensor distance is larger there than for deeper brain structures. In line with these observations, OPMs outperformed SQUIDs for two of the four patients with IEDs, with either a higher spike yield, or an ability to detect IEDs that were not observable in the SQUID data. For three patients the SNRs of IEDs were (slightly) lower in the OPM data than in the SQUID data, but with negligible effect sizes for two of these patients. Recording along two axes with the OPMs could be a contributing factor here, as this results in a slight reduction in sensitivity (∼30% reported by Osborne et al. (2018), but less by Rea et al. (2022)). A further possible explanation for this finding could be an insufficient coverage by the limited number of OPMs used in this study. This would particularly be an issue when an extended epileptogenic network is involved, which clinical MEG and sEEG recordings had previously shown for patient #1 (Table 1). That is, some of the IEDs in the extended network, or their field extrema, could have been missed by the small number of OPMs: the field maps for on-scalp sensors are much more confined than for SQUID-based systems due to the reduced source-sensor distance (see e.g. Boto et al. (2016)), such that IEDs’ sensor-level signatures can be easily missed, or be reduced in amplitude, when OPMs are not optimally placed. In other words, on-scalp OPM arrays have more focal fields-of-view (Iivanainen et al., 2017), implying that a more exact placement with respect to the underlying sources is required. It also means that with equal SNR, OPMs still offer higher spatial resolution than SQUIDS. Another explanation could be in the origin of the IEDs, which in patient #1 was in mesial temporal structures, for which the decreased source-sensor distance does not fully outweigh the decreased sensitivity of the OPMs compared to the SQUIDS (Boto et al., 2016). This could be remedied in future recordings by increasing the number of OPMs, including placement of OPMs over basal areas, and/or against the roof of the mouth (Tierney et al., 2021b).

Somewhat surprisingly, given the superficial origin of the IEDs, the average SNR of IEDs in patient #3 was comparable between the two systems. However, the SNR of the IEDs was low for both systems, and more IEDs were observed with the OPMs than with the SQUIDS. The inclusion of more, low-amplitude, IEDs reduced the average SNR for the IEDs in the OPM data. This would lead to the conclusion that the OPMs actually outperformed the SQUIDS, by their ability to detect such weak IEDs. However, sensor placement also had a big effect on the detectability of IEDs in this patient, as displacement of one OPM by 2.8 cm (Figure S2) made the IEDs undetectable. Another explanation for the findings could therefore be that the SNR was, also in this patient, reduced by sub-optimal placement of the OPMs. This is also the most likely explanation for the observed lower SNR for OPMs compared to SQUIDs for patient #4, who also had IEDs with a superficial origin (Table 1, Figure 3). It should be noted that even though the SNR differed significantly for this patient, due to the large number of IEDs, the actual difference in SNR was small (i.e. within the standard deviation of the SNR for the individual systems), with a medium effect size.

For patient #4, the spike yield was lower for the OPM data than for the SQUID data. This could potentially be explained by state-changes, as the OPM data were recorded in the morning and the SQUID data after lunch. This, in combination with the sleep-deprivation and the difference in patient-positioning (seated versus supine), resulted in a notable increase in drowsiness in the SQUID-session, which may have affected the yield of MEG abnormalities (Leach et al., 2006). Similarly, within a single session the IED yield could vary over time as well: for patient #3, for example, the first two (out of four) SQUID-recordings contained most of the IEDs. Unfortunately, the size of the OPMs, as well as the proximity of the cold-head, did not allow for a simultaneous recording with both systems. The spike yield could also have been artificially lowered in the OPM data due to movements. Movement-artefacts may have obscured IEDs during periods of movement. The patients had more freedom to move during the OPM recordings than during the (supine) SQUID recordings.

Unexpectedly, no IEDs could be found for patients #2 and #5 with either system. Patient #2 showed clear IEDs in both temporal lobes in the clinical MEG data that were recorded two and a half years prior to the current recordings, as well as in the stereo-EEG from 2 years ago. After the stereo-EEG the seizure frequency reduced, and the last seizure was a year ago, which may explain the absence of interictal activity in the current recordings. In the clinical MEG recordings of patient #5 from nearly 4 years ago there were clear spikes and polyspikes, and seizures remained, hence we do not have an explanation for the absence of IEDs in the current recordings.

### Seizure recordings

We demonstrated that seizure activity can be successfully recorded. The wearability of the sensors opens up the possibility for long-term observations, although to capture seizures in unselected patients one often has to record continuously for many days, as is done in an Epilepsy Monitoring Unit. Currently, even with a wearable OPM-based device, the recordings are restricted to the MSR due to the limited dynamic range of the OPMs, which limits the total length of a recording session. The dynamic range can be increased though by operating the OPMs in closed-loop (Fourcault et al., 2021), and by using alternative OPMs that do not rely on near-zero fields and that have a much larger dynamic range. Such sensors have already shown promise for the recording of MEG activity, even in an unshielded environment (Zhang et al., 2020). These developments could ultimately lead to long-term wearable MEG recordings in an unshielded environment, potentially removing the need for invasive stereo-EEG recordings.

### Future perspectives: increasing signal quality

Ictal MEG with SQUID-based systems is feasible for selected patients (see Stefan and Rampp (2020) for a review), namely those without (hyperkinetic) movements during the onset of their seizures. With OPMs the fixation to the scalp means that the movement restrictions are less severe, although movement of the OPMs through the remnant field-gradients still induces movement-artefacts (Figure 5). However, as long as the OPMs stay within their dynamic range so that the neuronal activity is captured alongside the artefacts, there is the possibility of recovering the signals of interest. We envisage that the increased spatial sampling with whole-head OPM coverage can be leveraged to achieve this, for example using beamforming. Seymour and colleagues have recently shown that straightforward pre-processing, involving the regression of motion-captured movement-parameters from OPM-recordings (removing artefacts

<0.5 Hz) and HFC (effectively the removal of the lower order external noise terms from SSS, removing artefacts 0-10 Hz), in combination with beamforming, allowed for movements of at least 1 meter (Seymour et al., 2021). It may be possible to improve on this by generating a model of the remnant fields inside the MSR, and using this model to predict and remove movement-related artefacts (Mellor et al., 2022). Moreover, reducing the field gradients in custom-designed MSRs with degaussing coils (Altarev et al., 2015), or by using field-nulling ‘matrix-coils’ that allow for accurate field control in a large volume (Holmes et al., 2021), would reduce the amplitude of movement-related artefacts to begin with. Finally, closed-loop designs can be used to further reduce movement-related artefacts, either at the OPM-level using a feedback signal to drive the on-board coils (requiring tri-axial sensitivity (Rea et al., 2022)), or through the field-nulling coils by ‘real-time’ updating of the coil calibration matrix, which would ensure optimal field cancellation during movements. With such technical developments, the increased SNR of OPMs (Boto et al., 2016; Boto et al., 2017; Iivanainen et al., 2017) could be utilised to its full extent to accurately localise the SOZ non-invasively. A key attraction of OPMs compared to SQUID systems is the flexibility of sensor placement, and the ability to detect and localise the SOZ in the (mesial) temporal lobe with OPMs could be further increased by strategic placement of the sensors (Tierney et al., 2021b).

### Future perspectives: helmet design

Bespoke rigid sensor arrays were created for all patients in this study, based on the individual anatomical MRI, with sensor placement based on the field maps of IEDs that had been identified in the previously recorded clinical MEG, in combination with information from stereo-EEG recordings (in the adult patients). However, no further optimisation of sensor placement was performed (Beltrachini et al., 2021; Iivanainen et al., 2021; Riaz et al., 2017). Particularly for systems with a limited number of sensors, the exact location of the sensors with respect to a region of interest becomes more important (Iivanainen et al., 2021). As mentioned above, the field maps generated for on-scalp sensors are quite compact, and local dense non-uniform sampling yields more information than uniform sampling of a larger area (Iivanainen et al., 2021). In future clinical studies, strong prior information about the expected generators of IEDs and ictal activity may not be available, rendering such an approach impractical. However, we envisage that multi-channel OPM-based systems with 50+ sensors and whole-head coverage (Hill et al., 2022) will become the norm. For such systems, uniform sampling may be sufficient, as long as the sensor spacing is approximately equal to the distance of the sensors to the closest source (Iivanainen et al., 2021). When using triaxial sensors, 75-100 uniformly placed sensors would provide sufficient spatial sampling (Tierney et al., 2022). It is still an open question what the optimal helmet design would be for larger clinical studies. Bespoke 3D-printed helmets are optimal in the sense that the sensors are fixed at known locations and with known orientations, yet their construction is time-consuming and costly. Flexible caps on the other hand can be re-used, but recordings may be more noisy due to sensor movements/rotations, and the exact sensor locations and orientations need to be determined. A compromise may be the use of a limited number of different-sized rigid helmets, with holders in which OPMs can be pressed onto the scalp (Zetter et al., 2019). The depth of the OPMs within their holder could be measured manually, or sensor positions and orientations could be determined automatically, using either an optical approach (Gu et al., 2021) or utilising the field-nulling coils (Iivanainen et al., 2022). The latter approach has the advantage that any changes in the orientations of the OPM’s sensitive axes due to crosstalk from neighbouring OPMs are already taken into account. These automatic approaches could also be used in combination with a design consisting of a bespoke rigid base to which a generic flexible cap with OPM-holders can be attached (https://quspin.com/experimental-meg-cap/).

### Limitations on the number of OPMs

An obvious limitation of this proof-of-principal study is the limited number of OPM sensors. By including well-characterised patients who had already undergone a successful clinical MEG that revealed IEDs that could be localised, as well as stereo-EEG (for the adult patients), we were able to increase the chances of capturing IEDs through strategic placement of the OPMs. However, despite this imbalance in the number of sensors (6 sensors/12 channels versus 102 sensors/306 channels), the OPM system performed as well as, if not better than, the SQUID-based system in terms of IED yield and/or SNR.

### Limitations on differences in recording conditions and data processing

Other factors that could have affected the direct comparison between the two systems include differences in pre-processing (HFC versus SSS), head modelling (single shell versus single sphere) and beamformer implementation (DAiSS versus Elekta beamformer), as well as the differences between recording sessions mentioned above (time-of-day, seated versus supine).

### Limitations on identification of IEDs

Interictal epileptiform discharges were primarily identified on the basis of visual inspection by an experienced EEG/MEG technician. A straightforward automatic algorithm that identified brief, sharp events that clearly stood out from the baseline (i.e. had a high Z-score), was used to identify other potential IEDs that were missed on visual inspection. Due to its simplicity, our automatic detector gave many false positives when the Z-score threshold was chosen such that (most) true positives were not missed, and careful visual assessment of the identified potential IEDs was therefore still required. For example, in the sensor-level SQUID data the algorithm initially locked-on to a strong ECG artefact that was present in some channels in some of the patients. This problem was mitigated by using only a sub-selection of channels for the automatic IED identification. Similarly, some IEDs that had been visually identified were missed by the automatic detector (false negatives) because of the IED-morphology. The data from patient #1 in particular contained polyspikes/spike-wave discharges that were sometimes missed by the detector, as were some small spikes for patient #3. Use of more sophisticated algorithms for the identification of interictal abnormalities (e.g. da Silva Lourenço et al. (2021); Kural et al. (2022)) would ease the objective comparison of the performance of both MEG systems, yet this was beyond the scope of the current manuscript.

A recent study showed that with the aid of artificial intelligence it is even possible to detect hippocampal epileptiform activity in the scalp EEG (Abou Jaoude et al., 2022). Simultaneous recordings of OPM- and EEG-data would enable a direct comparison between the two modalities regarding their ability to identify hippocampal IEDs. Although this is feasible (Ru et al., 2022), it also provides considerable engineering challenges, such as placement of the sensors/electrodes and how they may affect each other. Except for patient #7, we did not perform a direct comparison between the OPM data and the EEG, stereo-EEG and/or MEG data that had been recorded previously, for several reasons: i) the interval between these recordings was considerable (from half a year to several years), during which aging, changes in medication, or other factors could have affected the interictal activity; ii) such a comparison, for example in terms of IED-yield, would have been biased, since OPM-placement was based on these previous recordings. It is therefore not surprising that the OPM-results, for those patients with epileptiform activity in their OPM data, were in agreement with earlier EEG, stereo-EEG and/or MEG (see Table 1).

### Limitations on the statistical approach

For the statistical comparison of the SNR of the IEDs as obtained with the OPMs and SQUIDs a Mann-Whitney U-test was used. Hence, we used the assumption that the observations between the separate recordings were independent (since the recordings were performed at different times, and the IEDs themselves could therefore have changed over time). This is a conservative approach though, and one could increase statistical power by taking into account that the within-patient observations were not completely independent, for example through mixed-effects modelling (DeHart and Kaplan, 2019).

## 5. Conclusions

We have shown that interictal epileptiform activity can be reliably recorded with OPM-MEG, both in adults and paediatric populations. Moreover, the wearability of the sensors allowed for seizure recordings, even in the presence of significant movement. Overall, OPM data were very much comparable to those obtained with a cryogenic system, despite a potential lowering of the SNR of the IEDs due to suboptimal placement of the limited number of sensors. The relatively low cost of this technology, in combination with its reduced running and maintenance costs, means that OPM-based MEG could be used more widely than is the case with current MEG systems, and it may become an affordable alternative to scalp EEG, with the potential benefits of increased spatial accuracy, reduced sensitivity to volume conduction/field spread, and increased sensitivity to deep sources such as the hippocampus. Given its patient-friendliness, we envisage that wearable MEG will in the near future not only be used for presurgical evaluation of epilepsy patients, but also for diagnosis after a first seizure.

## Data Availability

Data and user-developed codes are available upon reasonable request to the corresponding author under the condition of an existing collaboration agreement.

## 6. Acknowledgements

We acknowledge the valuable contribution to this study by: the participants, and their parents, for taking part; Ilse van Straaten, Hanneke Ronner, Maeike Zijlmans, Kees Braun, and Floor Jansen for help with patient inclusion; the Development Team at the VUmc for construction of the field-nulling coils, Petteri Laine for construction of the cold-head compensation coils, and Mika Pajula and Rasmus Zetter for development of the filter unit for the cold-head compensation coils; Matthijs Grimbergen for development of the OPM-helmets; Annetje Guédon and Matthijs Grimbergen for development of system validation protocols; Peterjan Ris, Hans van der Horst, Theo Hillebrand, Prejaas Tewarie, James Osborne, and Vishal Shah for technical assistance; Anne van Nifterick, Francesco Galmozzi, Ida Nissen, Deborah Schoonhoven, Lennard Boon, Nandy Zwagerman, Nienke Scheltens, Nico Akemann, and Peterjan Ris for wiring of the field-nulling coils.

## 10. CRediT authorship contribution statement

**Arjan Hillebrand:** Conceptualization, Data curation, Formal analysis, Funding acquisition, Investigation, Methodology, Project administration, Resources, Software, Supervision, Validation, Visualization, Writing - original draft. **Niall Holmes:** Methodology, Resources, Software, Writing - review & editing. **Ndedi Sijsma:** Investigation. **George C. O’Neill:** Methodology, Software, Writing - review & editing. **Tim M. Tierney:** Methodology, Software, Writing - review & editing. **Niels Liberton:** Methodology, Resources. **Anine H. Stam:** Resources, Validation, Writing - review & editing. **Nicole van Klink:** Funding acquisition, Resources, Validation, Writing - review & editing. **Cornelis J. Stam:** Funding acquisition, Methodology, Resources, Software, Writing - review & editing. **Richard Bowtell:** Funding acquisition, Writing - review & editing. **Matthew J. Brookes:** Funding acquisition, Writing - review & editing. **Gareth R. Barnes:** Conceptualization, Funding acquisition, Methodology, Writing - review & editing.

