## Supplemental Material for "Improved non-invasive detection of ictal and interictal epileptiform activity using Optically Pumped Magnetometers"

### Semiology during video-EEG and OPM recording – Patient #7

During the video-EEG recordings, two types of seizure were recorded:

***Type 1***

Patient opens eyes, sits up if he was in supine position. For seizures that start when he is awake: he tries to say that he has a seizure, but does not finish sentence.

Looks frightened, eyes wide-open, face contorted. Left eye a bit more closed than right eye. Generalized increased tone for seconds (e.g. visible by raising of the shoulders), followed by hyperkinetic seizure with kicking, rocking movements with body, changing between sitting and lying down. Early in the seizure there is (if right-side is visible) a dystonia of the right hand. Large amplitude circular repetitive movement of the right arm. Eventually the movements stop, following which oromandibulaire automatisms (smacking/chewing), sometimes with right-sided manual automatisms, unforced slow head deviation to the left. Sometimes some noises/audible breathing. Impaired awareness. Post-ictally he speaks little and slowly, which may be aphasia, but is difficult to differentiate from a post-ictal state. During some seizures with initial hypermotor semiology as described above, this is not followed by temporal semiology, and patient also responds earlier when he is tested.

***Type 2***

Oromandibulaire automatisms, with some salivation, impaired awareness. Sometimes more subtle hyperkinetic semiology (subtle movements of legs/arms/trunk), generalized increased tone, blinking. Post-ictal possibly aphasia, but also difficult to differentiate from a post-ictal state (does not name, or very slow). On EEG these seizures have a right temporal onset and right temporal propagation pattern.

During the OPM recordings, the following semiology was observed:

Starts with eyes closed. Brief (~1 sec) blinking, then moves head forwards and back. Crosses arms in front of chest, looks up, frightened, with left eye closed and right eye wide open. Hypermobility, kicking, rocking movements with body and swinging left arm. Generalized increased muscle tone, unforced slow head deviation towards left, face contorted with mouth wide-open, following which some oromandibulaire automatisms (chewing). Early in the seizure there is a dystonia of the right hand. Eventually fewer movements, and leaning backward towards the left. Then sits straight and removes bandage from left arm and wipes nose with left arm. Impaired awareness. Remains restless for several minutes: head movements, intermitted eyes-open, fiddling with chin-strap, oromandibulaire automatisms, wipes nose with left arm.

**Conclusions**

The semiology observed during the OPM recording is in agreement with the semiology observed for seizures of Type 1 during video-EEG. sEEG has shown that these have a left temporal onset and propagation, or right frontal/temporal onset with rapid propagation to left temporal, with a further left temporal propagation (see also Table 1).

**Automatic identification of interictal epileptiform discharges**

Interictal epileptiform discharges (IEDs) were automatically identified using a matlab-implementation of the BrainSpike algorithm in BrainWave (version 0.9.162.4; developed by C.J.S.; available from https://home.kpn.nl/stam7883/). The algorithm detects spikes in sliding 1-sec windows, using steps of half a second. The windowed-data in a channel is converted to Z-scores, based on the mean and standard deviation for that window. If an event with a maximum Z-score exceeds a threshold within the middle 500 msec of the window (a threshold of 4, 3.5, 3.5, and 4.5 was used for patient #1, #3, #4, and #6, respectively), and if the event has a minimum and maximum duration (defined as the time between zero-crossings) of 20 and 200 msec, respectively, then it is marked as a potential IED. The algorithm was re-run on negated data in order to also identify 'negative' IEDs, and the IED with the maximum Z-score within a window, and across channels, was kept. Thus, a single Z-value was obtained for each IED, which was used as a proxy for the IED's SNR.

In order to reduce the number of false positives, the algorithm was run on only a selection of channels. The channel-selection was based on the field maps of the IEDs that had already been identified on the basis of a visual analysis of the data.

### Supplemental Figures

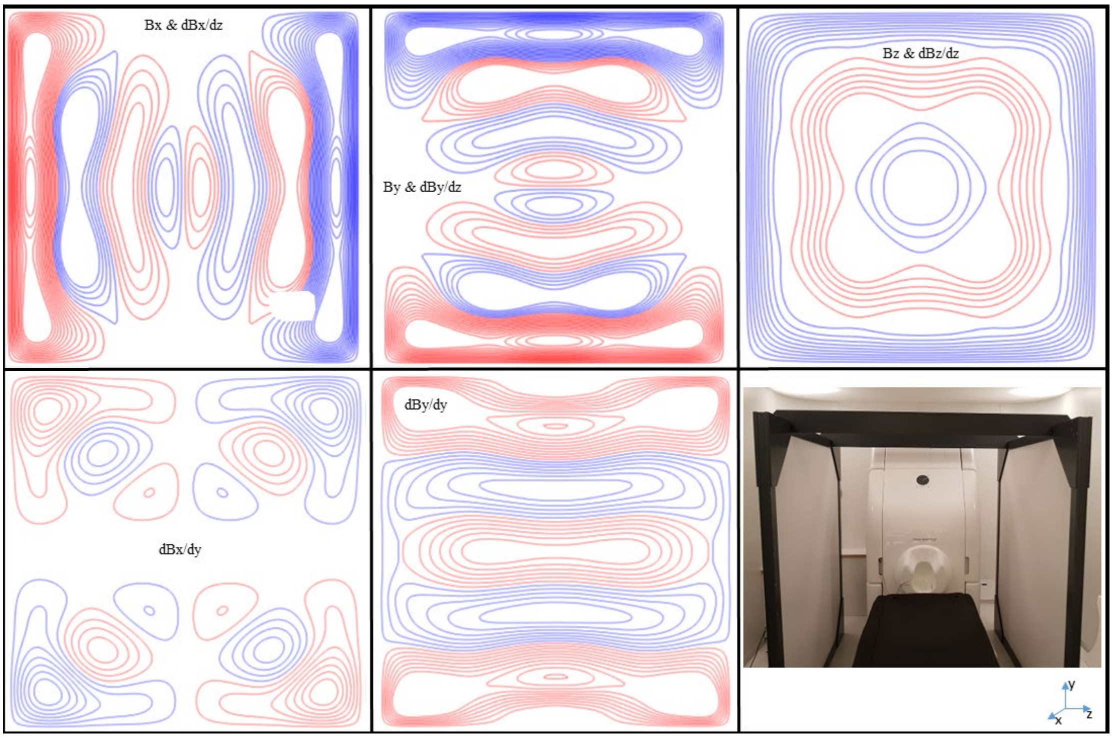


**Figure S1: Bi-planar nulling coils.** The natural symmetry shared by pairs of coils (Bx & dBx/dz, By & dBy/dz, and Bz & dBz/dz) was used in order to reduce the number of coils needed to minimise the field in all 3 directions, as well as the (linear) field gradients in the z-direction by a co-optimised coil design process (Holmes et al., 2019). Red and blue coloured wirepaths denote regions of opposing current flow. For the Bx & dBx/dz, By & dBy/dz coils driving the two planes with equal and opposite input currents generates a uniform magnetic field, driving the two planes with equal and same-signed currents generates a linear magnetic field gradient. For the Bz & dBz/dz coil, driving the two planes with equal and same-signed input currents generates a uniform magnetic field, driving the two planes with equal and opposite currents generates a linear magnetic field gradient. Two coils were added to allow nulling of the dBy/dy and dBx/dy gradients, in this case there is no shared symmetry which can be exploited so the windings of each plane were connected in anti-series. Grooves for each coil wirepath were routed into Forex PVC (polyvinylchloride) planes of 1.8 x 1.8 $m^2$. Enamelled copper wire of diameter 0.56mm was pressed into the grooves and fixed in place with tape at strategic positions. A front panel was added to prevent galvanic contact between the patients and the coils, and the stacked planes were placed inside a PVC frame (see lower-right sub-figure). The balanced coil pairs were connected individually, and the dBy/dy and dBx/dy coils in anti-series, to low-noise, 4 V, coil drivers (QuSpin Inc). Using this setup, the coils can generate uniform fields or field gradients within a 40 x 40 x 40 $cm^3$ volume between the centre of the panels. Bi-planar coil systems similar to those described here are commercially available (e.g. from Cerca Magnetics Limited, Nottingham, UK).

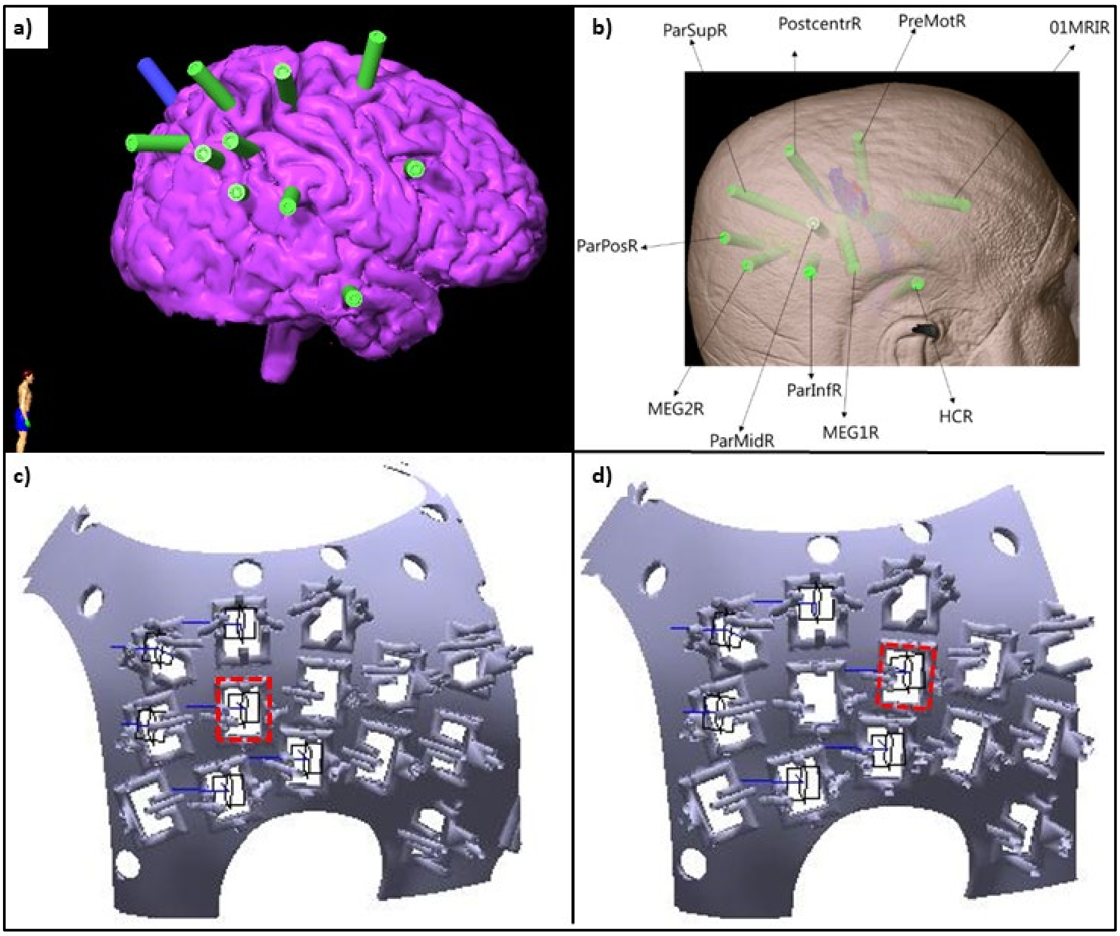


**Figure S2: sEEG setup and OPM-placement for patient #3.** Panels **a)** and **b)** show the depth-electrodes (green) with respect to the brain and scalp, respectively. The OPMs were placed over the right superior temporal/parietal lobe (electrode with label 'MEG1R') as this is where seizures started in the sEEG recordings, and where previous clinical MEG had localised IEDs. Using the OPM placement as in panel **c)**, the OPM that is indicated with a red rectangle recorded IEDs in the Bz channel. When this OPM was moved forward (panel **d)**), IEDs were not visible anymore. Blue arrows indicate the OPMs' sensitive axes (By and Bz), with By approximately parallel to the head and Bz approximately perpendicular to the head.

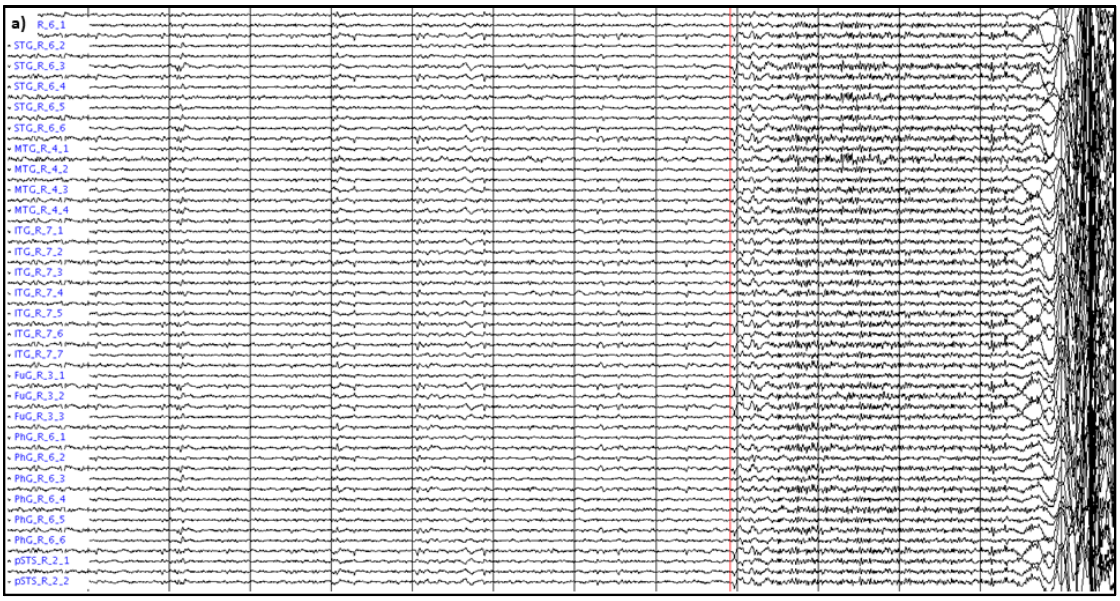
a)
R_6_1
STG_R_6_2
STG_R_6_3
STG_R_6_4
STG_R_6_5
STG_R_6_6
MTG_R_4_1
MTG_R_4_2
MTG_R_4_3
MTG_R_4_4
ITG_R_7_1
ITG_R_7_2
ITG_R_7_3
ITG_R_7_4
ITG_R_7_5
ITG_R_7_6
ITG_R_7_7
FuG_R_3_1
FuG_R_3_2
FuG_R_3_3
PhG_R_6_1
PhG_R_6_2
PhG_R_6_3
PhG_R_6_4
PhG_R_6_5
PhG_R_6_6
pSTS_R_2_1
pSTS_R_2_2

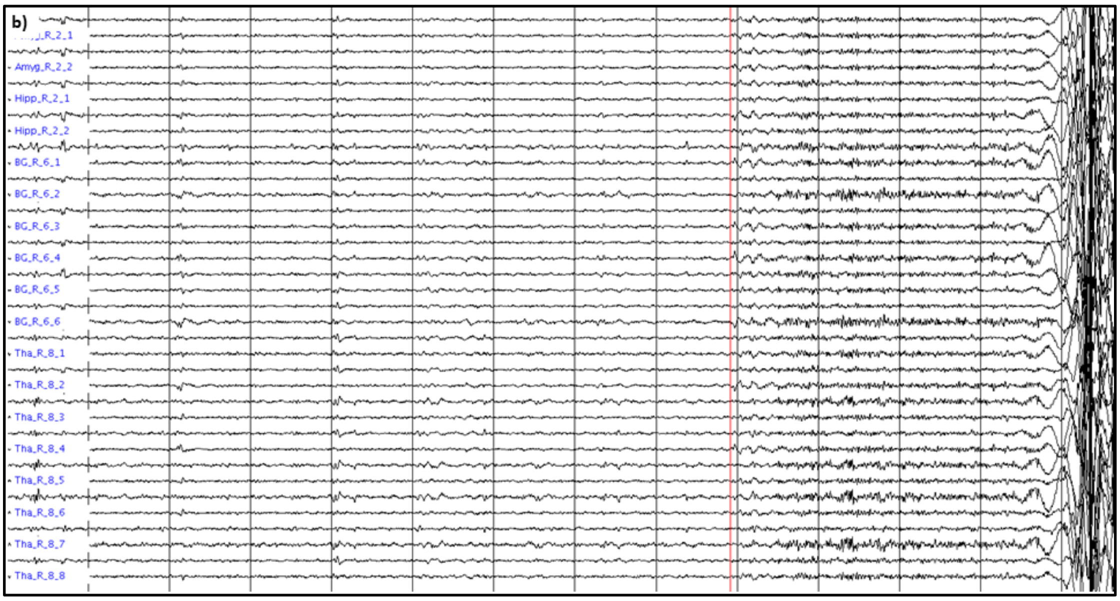
b)
R_2_1
Amyg_R_2_2
Hipp_R_2_1
Hipp_R_2_2
BG_R_6_1
BG_R_6_2
BG_R_6_3
BG_R_6_4
BG_R_6_5
BG_R_6_6
Tha_R_8_1
Tha_R_8_2
Tha_R_8_3
Tha_R_8_4
Tha_R_8_5
Tha_R_8_6
Tha_R_8_7
Tha_R_8_8

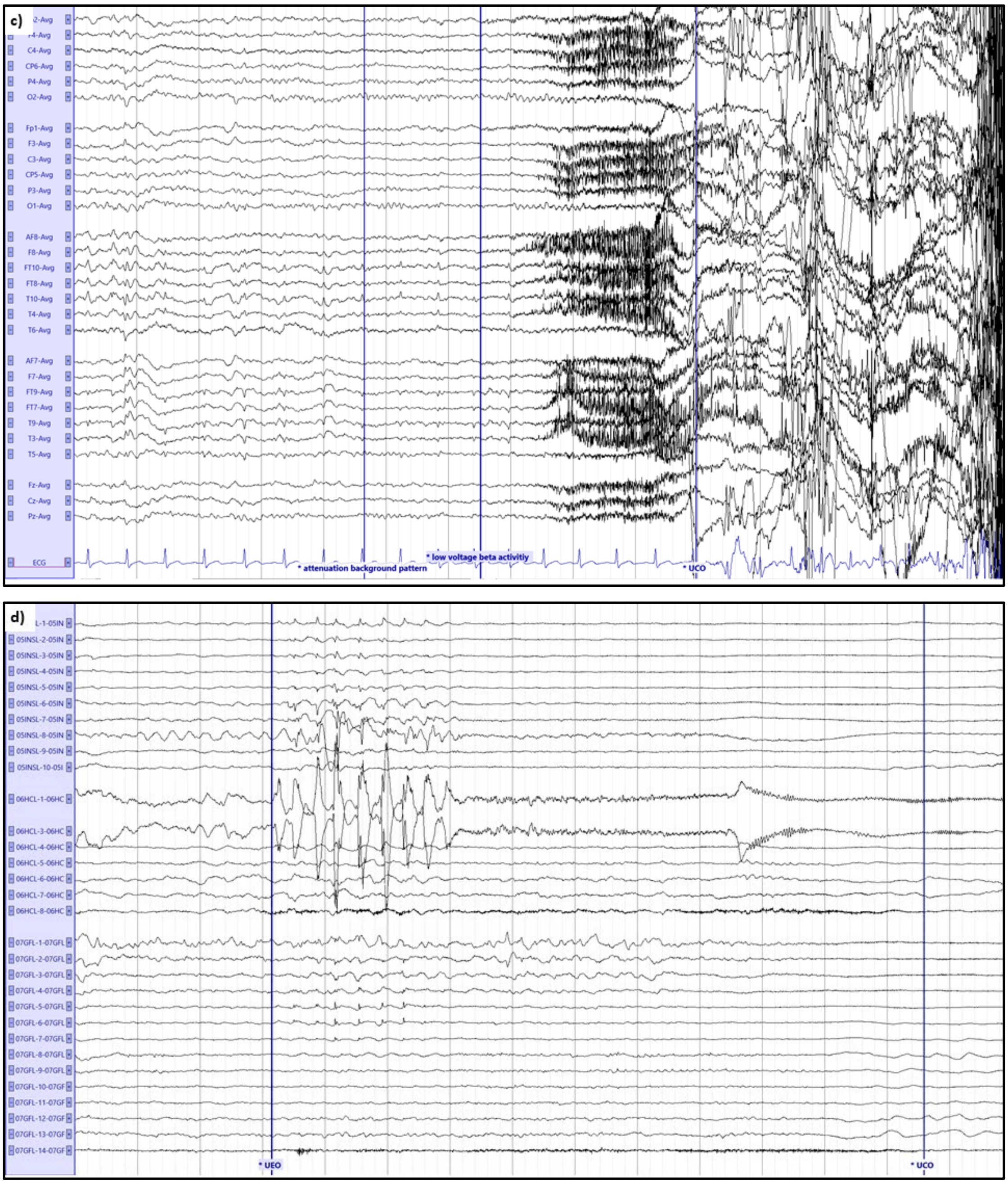


**Figure S3: Ictal onset for patient #7.** Source-reconstructed OPM data for temporal (**a**) and subcortical (**b**) channels in the BNA atlas (see (Fan et al., 2016) for nomenclature). Channels from left and right hemisphere are alternated. The grey vertical lines mark 1 second of data, that were filtered between 3 - 48 Hz. Note the increase in fast activity, simultaneously in both hemispheres, after about 9 secs (red vertical line), marking the start of the seizure. This is followed by artefacts due to movement during the seizure. Panel **c** shows a segment of seizure activity, Type 1, from a video-EEG recorded in 2021. Seizure from drowsy state, EEG applied according to the 10-20 system with extra electrodes from the 10-10 system (anterior frontal (AF7/8), frontotemporal (FT9/10, FT7/8), temporal (T9/10), centroparietal (CP5/6)). Average montage. The grey vertical lines mark 1 second of data, amplitude 70 µV/cm, high and low pass filter of 0.27 Hz and 15.0 Hz, respectively. The seizure starts with attenuation of the background pattern ($1^{st}$ blue vertical line from the left), followed with diffuse beta activity (2nd blue vertical line; mainly fronto-temporal), then beta mixed with muscle artefact (although no patient movements were visible yet), followed by large movement artefacts. Note the similarity in temporal evolution in the video-EEG and OPM recording. Panel **d** shows a segment of seizure activity, Type 1, from an sEEG recording from 2021. Only electrodes involved in the seizure onset are depicted (05INSL: entry: frontal, target: anterior insula. 06HCL: entry: temporal, target: left hippocampus. 07GFL: entry: temporal, target: left fusiform gyrus). Bipolar montage. The grey vertical lines mark 1 second of data, amplitude 700 µV/cm, high and low pass filter of 0.27 Hz and 100 Hz, respectively. UEO: unequivocal electrographic onset. UCO: unequivocal clinical onset. Seizure onset (1st blue vertical line): 2.5 Hz spike-wave complexes on position 6HCL1-3, also visible at 5INSL1-8 and less pronounced at 7GFL1-8, followed by irregular high frequency activity and subsequent beta activity with a frequency of ~20 Hz on 06HCL1-3. Seizure propagation (not shown) in consecutive order: left posterior temporal lobe (GFL), left anterior cingulate cortex (anterior and dorsal part) and left frontobasal area.

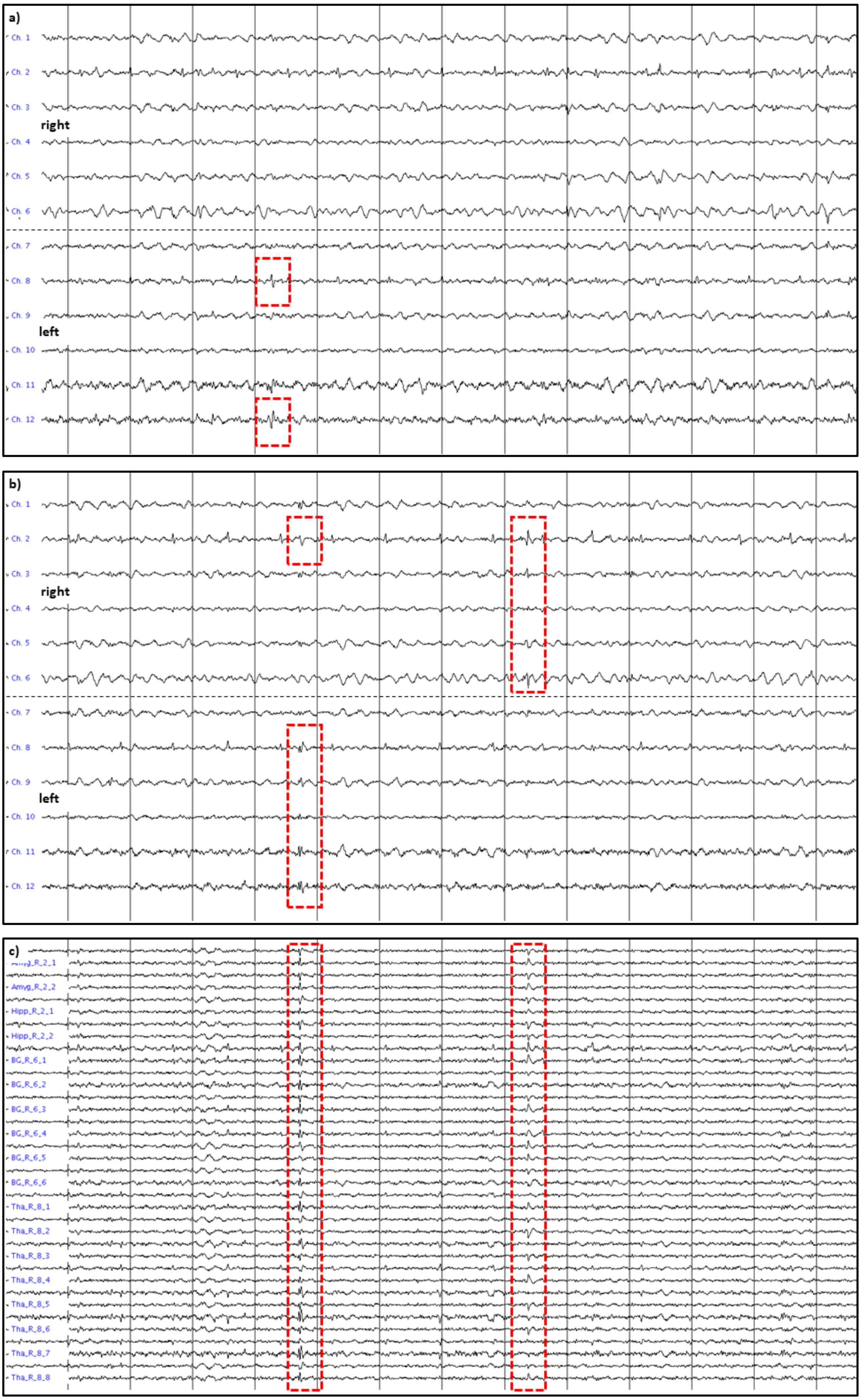
a)
Ch. 1
Ch. 2
Ch. 3
right
Ch. 4
Ch. 5
Ch. 6
Ch. 7
Ch. 8
Ch. 9
left
Ch. 10
Ch. 11
Ch. 12
b)
Ch. 1
Ch. 2
Ch. 3
right
Ch. 4
Ch. 5
Ch. 6
Ch. 7
Ch. 8
Ch. 9
left
Ch. 10
Ch. 11
Ch. 12
c)
Amyg_R_2_2
Hipp_R_2_1
Hipp_R_2_2
BG_R_6_1
BG_R_6_2
BG_R_6_3
BG_R_6_4
BG_R_6_5
BG_R_6_6
Tha_R_8_1
Tha_R_8_2
Tha_R_8_3
Tha_R_8_4
Tha_R_8_5
Tha_R_8_6
Tha_R_8_7
Tha_R_8_8

**Figure S4: Interictal activity for patient #7.** Independent IEDs were observed over the left (**a**) and right (**b**) temporal lobes, as well as simultaneously over both temporal lobes (**b**). The first 6 channels were placed over the right hemisphere and the last 6 channels over the left hemisphere; alternating channels show recording in the OPMs' By and Bz direction. Note the ECG artefact on channel 2 and 8, which was removed/reduced through beamforming (**c**). Panel **c** shows the same segment of data as in panel **b**, source-reconstructed to the subcortical channels in the BNA atlas. Channels from left and right hemisphere are alternated. The grey vertical lines mark 1 second of data, that were filtered between 3 - 48 Hz.
